# Plasma proteomic markers of cognitive and brain aging in a multi-ethnic cohort

**DOI:** 10.64898/2026.11.22.26368682

**Authors:** Brendan Wood, Madeline Wood Alexander, Andrew Clappison, Katie L. Vandeloo, Rachel Yep, Rowan Saloner, Joshua Downer, Kaitlin B. Casaletto, Michelle Caunca, Rohina Kumar, Simran Malhotra, Angelina Zhang, Tulip Marawi, Valery Sit, Ella Kim, Alexander J. Nyman, Tallinn F. L. Splinter, Min Su Kang, Walter Swardfager, Thomas K. Karikari, Sandra E. Black, Julie Ottoy, Maged Goubran, Jennifer S. Rabin, the Alzheimer’s Disease Neuroimaging Initiative

**Affiliations:** Hurvitz Brain Sciences Program, Sunnybrook Research Institute, Toronto, Ontario, Canada; Department of Medical Biophysics, University of Toronto, Toronto, ON, Canada; Rehabilitation Sciences Institute, Temerty Faculty of Medicine, University of Toronto, Toronto, Ontario, Canada; Harquail Centre for Neuromodulation, Sunnybrook Health Sciences Centre, University of Toronto, Toronto, Ontario, Canada; Ann S. Bowers Women’s Brain Health Initiative, University of California, California, USA; Department of Psychological Clinical Sciences, University of Toronto Scarborough, Ontario, Canada; Fein Memory and Aging Center, Department of Neurology, Weill Institute for Neurosciences, University of California, San Francisco, CA, USA; Neurovascular Division, Department of Neurology, University of California, San Francisco, San Francisco, CA, USA; Department of Neurology, University of Wisconsin-Madison School of Medicine and Public Health, Madison, WI, USA; Department of Pharmacology & Toxicology, University of Toronto, Ontario, Canada; Black Centre for Brain Resilience & Recovery, Hurvitz Brain Sciences Program, Sunnybrook Research Institute, University of Toronto, Toronto, ON, Canada; Department of Psychiatry, School of Medicine, University of Pittsburgh, Pittsburgh, PA, USA; Biofluid Biomarker Laboratory, Western Psychiatric Hospital, University of Pittsburgh Medical Center, Pittsburgh, PA, USA; Alzheimer’s Disease Research Center, University of Pittsburgh, Pittsburgh PA, USA; Clinical and Translational Science Institute, University of Pittsburgh, Pittsburgh, PA, USA; Temerty Faculty of Medicine, Department of Medicine (Neurology), University of Toronto, Toronto, ON, Canada; Physical Sciences Platform, Sunnybrook Research Institute, Toronto, Ontario, Canada

## Abstract

Advances in ultrasensitive proteomics offer opportunities to characterize molecular pathways underlying brain aging. We measured 220 plasma proteins using NULISAseq Neuro220 in 250 Chinese, South Asian, and non-Hispanic White older adults (mean age=68.2 years; 70.4% female). Proteins implicated in neurodegenerative diseases (including brain-derived phosphorylated Tau; BDpTau), synaptic function, oxidative stress, and inflammation were associated with poorer cognition. Phosphorylated Tau205 was associated with reduced entorhinal thickness, whereas inflammatory proteins were associated with imaging markers of cerebral small vessel disease. Several findings were replicated in 1,381 participants in the Alzheimer’s Disease Neuroimaging Initiative, including associations of BDpTau species, NPTX2, BACE1, TREM1, CCL17, PRDX6, and IGF1R with cognitive decline and clinical progression. Cardiometabolic risk factors were associated with widespread proteomic alterations, many of which were also linked to poorer brain health. These results identify promising molecular markers of brain aging and extend proteomic research to ethnically diverse populations historically underrepresented in Alzheimer’s disease research.

## 1. INTRODUCTION

Cognitive and brain aging are shaped by multiple, co-occurring biological processes, including neurodegenerative pathology, cerebrovascular dysfunction, inflammation, metabolic dysregulation, and synaptic loss.^1–4^ Identifying the molecular signatures of these processes may help identify individuals at increased risk of cognitive impairment and reveal potential targets for intervention.^5–8^ The plasma proteome offers a minimally invasive means of investigating these processes.^9^ Despite major advances in blood biomarkers for Alzheimer’s disease (AD),^10,11^ our understanding of the broader plasma proteomic signatures associated with cognitive and brain aging remains limited, particularly in ethnically diverse populations.

Recent advances in high-sensitivity multiplex proteomics now enable the simultaneous quantification of hundreds of circulating proteins relevant to brain aging and neurological disease.^12^ Studies using the NUcleic acid Linked Immuno-Sandwich Assay (NULISAseq) CNS 120 platform have identified molecular signatures associated with cognition, neuroimaging measures, and AD pathology, spanning inflammatory, vascular, synaptic, and neurodegenerative pathways.^13–16^ The expanded NULISAseq Neuro 220 panel includes approximately 100 additional brain-relevant proteins, including phosphorylated tau species beyond pTau217 (e.g., pTau205 and pTau212^17–21^), brain-derived pTau species, and additional inflammatory and synaptic targets. Whether these expanded targets provide further insight into cognitive and brain aging remains largely unexplored.

Another important limitation of dementia biomarker research is the underrepresentation of ethnically diverse populations. This raises questions about whether previously identified proteomic associations generalize across populations.^22,23^ This is particularly relevant given population differences in cardiometabolic health, including hypertension and diabetes, which may influence circulating protein profiles and contribute to cognitive impairment and cerebrovascular disease.^24–26^ Social and structural determinants of health may further affect these relationships.^27^ Examining whether cardiometabolic risk factors and brain aging share circulating proteomic signatures may provide insight into the biological processes connecting systemic and brain health. However, these relationships remain poorly characterized in ethnically diverse aging populations.

Here, we used the NULISAseq Neuro 220 panel to characterize circulating proteomic signatures of cognitive, brain, and cardiometabolic health in the Canadian Multi-Ethnic Research on Aging (CAMERA) study. This deeply phenotyped cohort includes adults aged 55 to 85, primarily from Chinese, South Asian, and non-Hispanic White backgrounds. Our primary objective was to characterize these associations across the full cohort, extending the investigation of plasma proteomic signatures of brain aging to populations historically underrepresented in dementia biomarker research. We identified proteins associated with cognitive performance and MRI markers of neurodegeneration and cerebral small vessel disease, spanning processes related to AD pathobiology, synaptic function, inflammation, and oxidative stress. Several proteomic associations were replicated in the Alzheimer’s Disease Neuroimaging Initiative (ADNI), an independent observational cohort with substantially different demographic and clinical characteristics. We further identified overlapping proteomic signatures of cardiometabolic risk and adverse cognitive and neuroimaging outcomes, particularly involving inflammatory proteins. Together, these findings broaden our understanding of the circulating proteome of cognitive and brain aging and identify biological processes shared with cardiometabolic health

## 2. RESULTS

### 2.1. Participant characteristics

We included cross-sectional data from 250 adults in the CAMERA study.^23^ Participants included 82 Chinese, 74 South Asian, 83 non-Hispanic White, and 11 individuals of other ethnicities (Table 1; Figure 1A). The mean (*SD*) age was 68.2 (6.78) years and 29.6 % (*N*=74) were male. Age, sex, and education did not differ significantly across ethnic groups (Figs. 1B-1C). Fifty-nine participants (23.6%) were *APOE* ε*4* carriers, with a higher prevalence among non-Hispanic White than Chinese participants (33.3% vs. 14.5%; Fig 1D). Most participants were cognitively unimpaired (Clinical Dementia Rating [CDR] global score = 0), with only six participants (2.4%) scoring 0.5.

**Figure 1.**
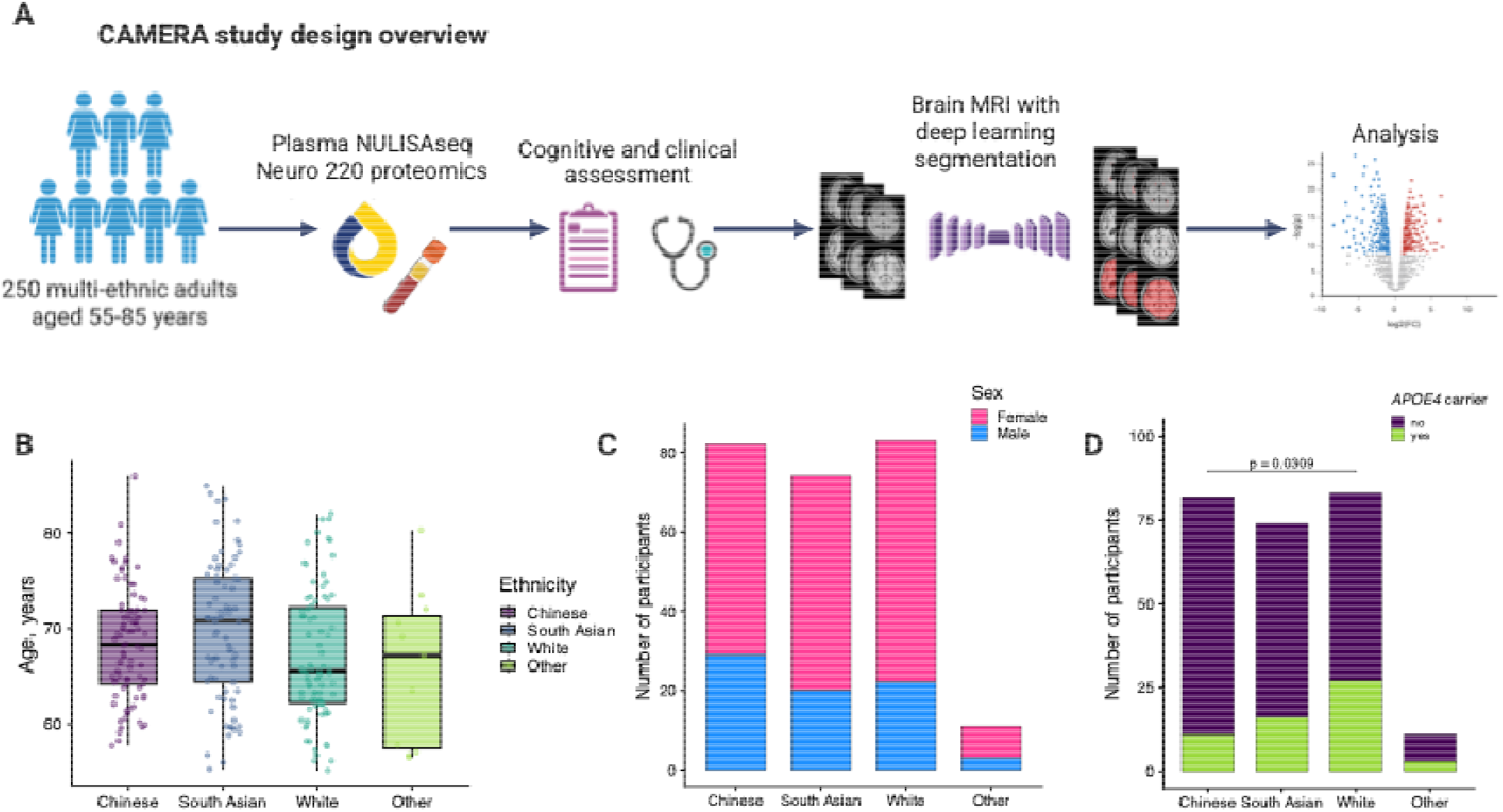
Study design and participant characteristics. **A.** Overview of present study, comprising 250 adults aged 55-85 from the Canadian Multi-Ethnic Research on Aging (CAMERA) cohort who underwent plasma proteomic profiling using the NULISAseq Neuro 220 panel, clinical and neuropsychological assessments, and brain MRI. **B.** Age and **C.** sex distributions across ethnic groups. **D.** *APOE4* ε4 carrier frequency across ethnic groups

**Table 1.** Demographic and clinical characteristics of participants in the Canadian Multi-Ethnic Research on Aging (CAMERA) cohort.

| **Variable** | **All participants (*N* = 250)** | **By ethnicity** |  |  |  |  |  |
|---|---|---|---|---|---|---|---|
|  |  | **Chinese (*N* = 82)** | **South Asian (*N* = 74)** | **White (*N* = 83)** | **Other\* (*N* = 11)** | **Overall p-value** | **Pairwise contrasts p-values** |
| Age, mean (*SD*) years | 68.20 (6.78) | 68.29 (5.82) | 69.75 (7.32) | 67.03 (6.80) | 65.85 (8.10) | .05 | - |
| Male sex, *N* (%) | 74 (29.6) | 29 (35.4) | 20 (27.0) | 22 (26.5) | 3 (27.3) | .58 | - |
| Education, mean (*SD*) years | 16.42 (2.24) | 16.20 (2.09) | 16.96 (2.35) | 16.28 (2.22) | 15.45 (2.21) | .06 | - |
| English as a first language (EFL), *N* (%) | 89 (35.6) | 8 (9.76) | 16 (21.6) | 62 (74.7) | 3 (27.3) | <.001 | SA vs. C: .15<br>SA vs. W: 1.3e-10<br>SA vs. O: .70<br>C vs. W: 4.4e-17<br>C vs. O: .24<br>W vs. O: .01 |
| *APOE ε4* carrier, *N* (%) | 57 (22.8) | 11 (13.4) | 16 (21.6) | 27 (32.5) | 3 (27.3) | .03 | SA vs. C: .83<br>SA vs. W: .76<br>SA vs. O: 1<br>C vs. W: .03<br>C vs. O: 1<br>W vs. O: 1 |
| CDR global score, *N* (%) |  |  |  |  |  | .35 | - |
| 0 | 244 (97.6) | 78 (95.0) | 73 (98.6) | 82 (98.8) | 11 (100) |  |  |
| 0.5 | 6 (2.4) | 4 (5.0) | 1 (1.4) | 1 (1.2) | 0 (0.0) |  |  |
| Born in Canada, *N* (%) | 75 (30.0) | 11 (13.9) | 1 (1.5) | 62 (74.7) | 1 (9.1) | <.001 | SA vs. C: .02<br>SA vs. W: 1.9e-22<br>SA vs. O: .52<br>C vs. W: 4.5e-15<br>C vs. O: 1<br>W vs. O: .0001 |
| Body mass index, mean (*SD*) | 25.25 (4.13) | 23.86 (3.79) | 25.60 (4.07) | 25.98 (4.11) | 27.76 (4.59) | .001 | SA vs. C: .03<br>SA vs. W: .56<br>SA vs. O: .49<br>C vs. W: .005<br>C vs. O: .08<br>W vs. O: .49 |
| HbA1c, mean (*SD*) % | 5.82 (0.52) | 5.98 (0.49) | 5.93 (0.63) | 5.55 (0.31) | 5.88 (0.37) | <.001 | SA vs. C: 1<br>SA vs. W: 4.9e-5<br>SA vs. O: 1<br>C vs. W: 2.4e-9<br>C vs. O: 1<br>W vs. O: .06 |
| HDL cholesterol, mean (*SD*) mmol/L | 1.74 (0.48) | 1.80 (0.47) | 1.62 (0.44) | 1.81 (0.51) | 1.48 (0.34) | .01 | SA vs. C: .07<br>SA vs. W: .07<br>SA vs. O: .49<br>C vs. W: .84<br>C vs. O: .07<br>W vs. O: .07 |
| LDL cholesterol, mean (*SD*) mmol/L | 2.87 (1.05) | 2.78 (1.08) | 2.66 (1.15) | 3.12 (0.88) | 3.06 (0.91) | .04 | SA vs. C: 1<br>SA vs. W: .04<br>SA vs. O: .82<br>C vs. W: .17<br>C vs. O: 1<br>W vs. O: 1 |
| Triglycerides, mean (*SD*) mmol/L | 1.12 (0.59) | 1.09 (0.62) | 1.29 (0.64) | 1.01 (0.50) | 1.16 (0.49) | .03 | SA vs. C: .27<br>SA vs. W: .02<br>SA vs. O: 1<br>C vs. W: 1<br>C vs. O: 1<br>W vs. O: 1 |
| Systolic blood pressure, mean (*SD*) mmHg | 131.48 (15.60) | 131.19 (15.85) | 132.61 (12.91) | 131.24 (17.69) | 127.57 (14.55) | .79 | - |
| Anti-hypertensive medications, *N* (%) | 77 (30.8) | 29 (35.4) | 26 (35.1) | 19 (22.9) | 3 (27.3) | .27 | - |
| Lipid-modifying medications, *N* (%) | 86 (34.4) | 30 (36.6) | 34 (45.9) | 17 (20.5) | 5 (45.5) | .007 | SA vs. C: .78<br>SA vs. W: .006<br>SA vs. O: 1<br>C vs. W: .13<br>C vs. O: 1<br>W vs. O: .49 |
| Glucose-lowering medications, *N* (%) | 28 (11.2) | 13 (15.9) | 12 (16.2) | 2 (2.4) | 1 (9.1) | .01 | SA vs. C: 1<br>SA vs. W: .02<br>SA vs. O: 1<br>C vs. W: .02<br>C vs. O: 1<br>W vs. O: 1 |

Overall p-values represent unadjusted comparisons across ethnic groups using analysis of variance for continuous variables and χ² for categorical variables, as appropriate. Where nominally significant overall ethnic differences were observed, Holm-corrected p-values are also presented for pairwise contrasts. Other ethnicities included 5 South East Asian, 1 Hispanic/Latino/Spanish, 1 Middle Eastern/North African, 3 mixed ethnicities, and 2 who preferred not to describe Abbreviations: APOE, apolipoprotein E; BMI, body mass index; CDR, Clinical Dementia Rating; HbA1c, glycated hemoglobin; HDL, high-density lipoprotein; LDL, low-density lipoprotein; SD, standard deviation; SA, South Asian; C, Chinese; W, White; O, Other.

### 2.2. Age, sex, and *APOE* ε*4* associations with plasma proteomic profiles

Given the established roles of age, sex, and *APOE* ε*4* carriage in brain aging and dementia,^3,28,29^ we first examined their associations with 200 NULISAseq proteins and three protein ratios reflecting AD-related amyloid and tau pathology (i.e., pTau217/Aβ42, pTau217/MAPT [pTau217%], brain derived [BD] pTau217/MAPT [BDpTau217%]).^20^

In multivariable linear models including terms for age, sex and ethnicity, age was positively associated with 67 proteins (50 FDR, 17 nominal) and negatively associated with 4 proteins (2 FDR-significant, 2 nominally significant; Table S1; Figure 2A). The strongest positive associations included proteins linked to neurodegenerative disease and neurodegeneration (NEFL, NEFH, BDpTau212), biological aging (GDF15, Figure 2B), inflammation (GFAP, TREM1, TREM2 CHI3L1), and cardiac stress (NTproBNP, Figure 2C). By contrast, extracellular matrix glycoprotein EFEMP and growth factor receptor EFGR were negatively associated with age.

**Figure 2.**
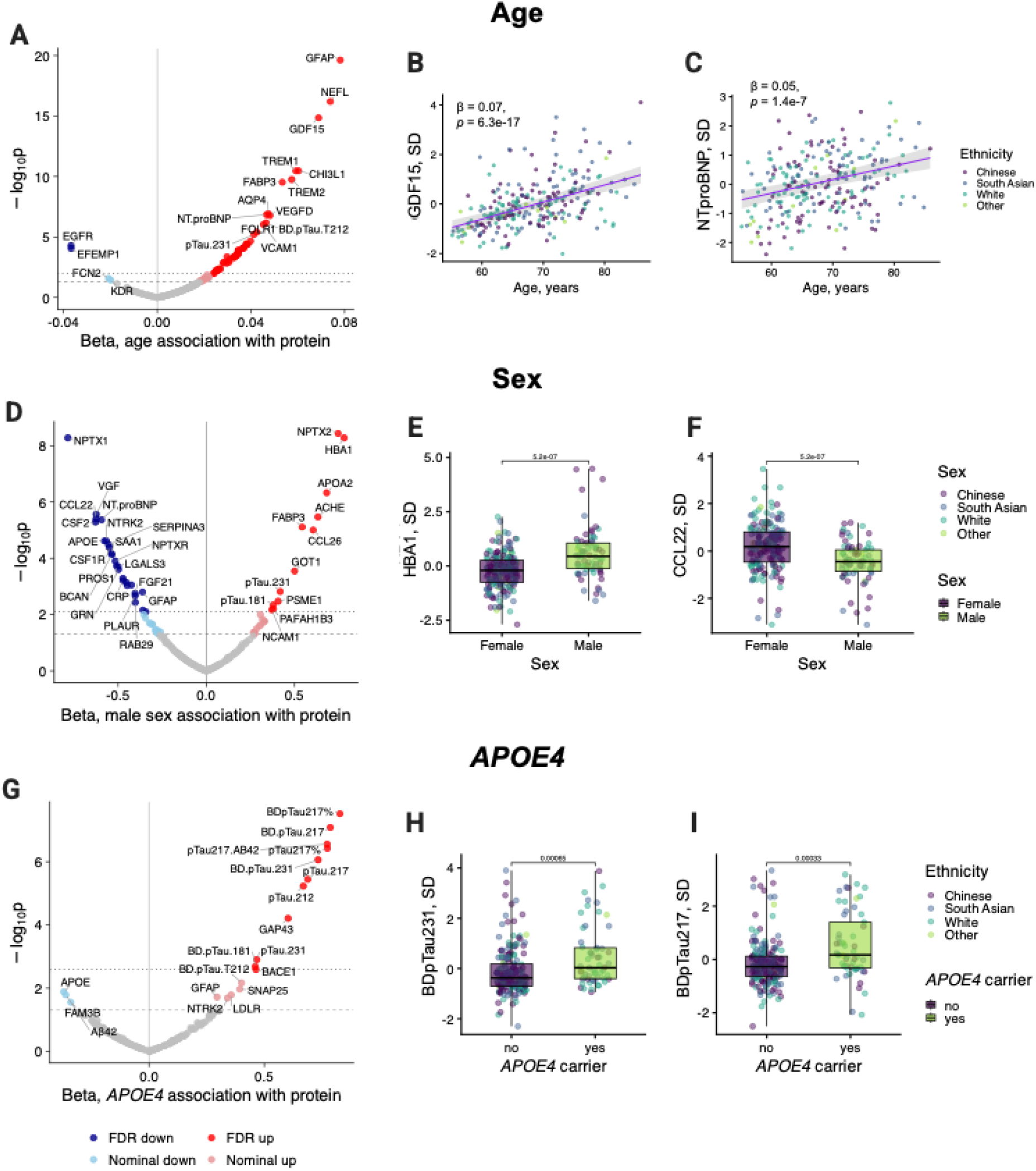
Age, sex and APOE ε4 genotype are associated with distinct plasma proteomic profiles. **A.** Associations of age with plasma proteins from linear models adjusted for sex and ethnicity. Older age was associated with higher levels of 67 proteins (50 FDR, 17 nominal) and lower levels of four proteins (2 FDR, 2 nominal). **B.** Association of age with GDF15, a canonical marker of biological aging. **C.** Association of age with NTproBNP, a marker of cardiac stress. **D.** Sex differences in plasma protein levels from linear models adjusted for age and ethnicity. Men had higher levels of 23 proteins (12 FDR, 11 nominal), whereas women had higher levels of 39 proteins (26 FDR, 13 nominal). **E.** Sex differences in HBA1 levels **F.** Sex differences in CCL22 levels. **G.** Associations of *APOE* ε4 carrier status with plasma proteins from linear models adjusted for age, sex and ethnicity. Carriers had higher levels of 16 proteins (11 FDR, 5 nominal) and lower levels of three proteins (all nominally significant) compared with non-carriers. **H.** Association of *APOE* ε4 carrier status with BDpTau231 levels. **I.** Association of *APOE* ε4 carrier status with BDpTau217 levels.

In the same models, 23 proteins were higher in men (12 FDR, 11 nominal) and 39 were higher in women (26 FDR, 13 nominal; Table S1; Figure 2D). Top proteins elevated in men included the alpha-globin subunit of hemoglobin HBA1 (Figure 2E) and the synaptic and partially testes-secreted protein NPTX2, whereas those elevated in women included the synaptic protein NPTX1, cardiac stress marker NTproBNP, and chemokine CCL22 (Figure 2F).

In models adjusted for age, sex, and ethnicity, *APOE* ε*4* carriers exhibited higher levels of 16 proteins (11 FDR, 5 nominal; Table S2; Figure 2G), including phosphorylated tau species BDpTau231 (Figure 2H), BDpTau217 (Figure 2I), pTau217, and pTau212, as well as the synaptic marker GAP43 and BACE1, which is involved in β-amyloid precursor protein processing. Conversely, *APOE* ε*4* carriers exhibited nominally lower levels of Aβ42 (reflecting less favorable amyloid levels), APOE, and the metabolic protein FAM3.

Given evidence that associations of *APOE* ε*4* with AD risk and biomarkers may differ across ancestral populations,^30^ we conducted exploratory analyses stratified by self-reported ethnicity (South Asian, Chinese, and non-Hispanic White; Table S2; Extended data Figure 1). *APOE* ε*4* carriers generally had higher levels of AD-related proteins across all three groups, including several p-tau species, BACE1, and GAP43. Effect estimates for pTau species tended to be larger among non-Hispanic White participants, whereas associations with BACE1 and GAP43 were more similar across groups. Exploratory analyses testing the interaction between ethnicity and *APOE* ε*4* carriage showed nominally significant interactions on several p-tau species (i.e., BDpTau181, pTau181, pTau212, among others), with stronger associations in White than South Asian and Chinese participants. By contrast, associations of *APOE* ε*4* with p-tau levels did not significantly differ between South Asian and Chinese individuals (Table S2; Extended data Figure 1).

### 2.3. Plasma proteins associated with cognition

We next examined associations of circulating proteins with cognitive performance and three MRI-derived measures of brain aging (Figure 3A). Cognition was assessed using a composite adapted from the Preclinical Alzheimer Cognitive Composite (PACC),^31^ comprising predominantly nonverbal tests to reduce language-related influences across ethnic groups. In multivariable linear models adjusted for age, sex, ethnicity, years of education, and English as a first language, 58 proteins were negatively associated with cognition (15 FDR, 43 nominal; Table S3; Figure 3B). The strongest associations with poorer cognitive performance included oxidative stress-related protein PRDX6, SNARE protein STX4, NPTX2, and several phosphorylated tau species including BDpTau181 and pTau217 (Figure 3C).

**Figure 3.**
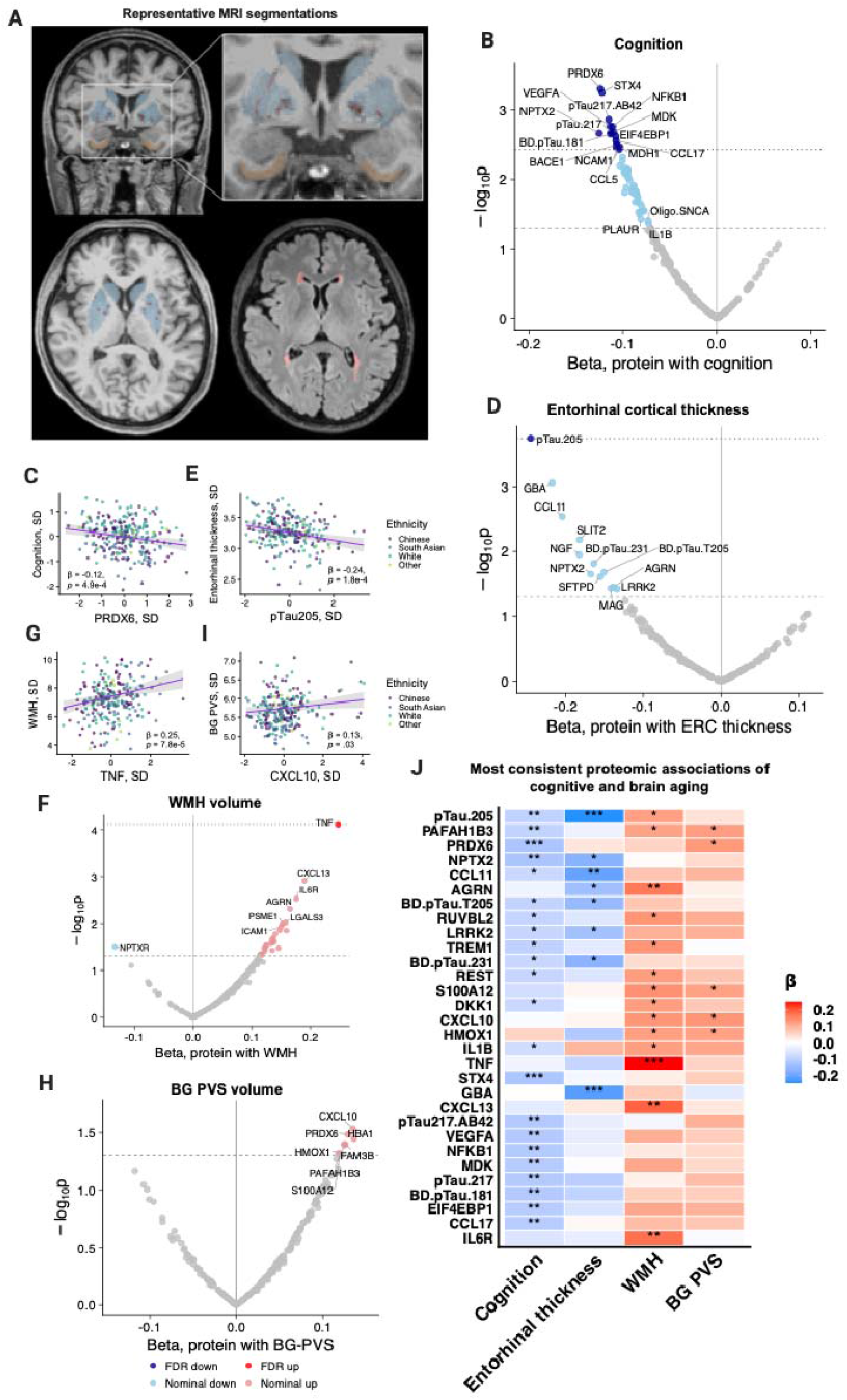
Plasma proteomic signatures of cognitive performance and MRI-derived measures of brain aging. **A.** Representative MRI segmentations of the entorhinal cortex (top, yellow), basal ganglia enlarged perivascular spaces (BG-PVS; BG in blue, PVS in red, top and bottom left) and white matter hyperintensities (WMH; bottom right, red). **B.** Associations of plasma proteins with cognition, adjusted for age, sex, ethnicity, years of education, and English as a first language. Fivty-eight proteins were negatively associated with poorer cognitive scores (15 FDR, 43 nominal). **C.** Association of the oxidative-stress marker PRDX6 with cognition. **D.** Associations of plasma proteins with entorhinal cortical thickness, adjusted for age, sex and ethnicity. Twelve proteins were negatively associated with entorhinal thickness (1 FDR, 11 nominal). **E.** Association of pTau205 with entorhinal thickness. **F.** Associations of plasma proteins with log-transformed WMH volume, adjusted for age, sex, ethnicity, and intracranial volume. Twenty-seven proteins were positively associated with greater WMH volume (1 FDR, 26 nominal). **G.** Association of TNF with WMH volume. **H.** Associations of plasma proteins with log-transformed BG-PVS volume, adjusted for age, sex, ethnicity, and intracranial volume. Seven proteins were positively associated with BG-PVS volume (all nominal). **I.** Association of CXCL10 with BG-PVS volume. **J.** Heatmap of the 30 proteins showing the most consistent associations across four cognitive and MRI outcomes selected and ordered according to the number of nominally significant associations and magnitude of effect estimates. The asterisks reflect nominal p-values. * *p* < .05, ** *p* < .01, *** *p* < .001.

### 2.4. Plasma proteins associated with entorhinal thickness

We next examined whether circulating proteins were associated with entorhinal cortical thickness, a region particularly vulnerable to aging and early AD-related neurodegeneration.^32^ Among 233 CAMERA participants with available brain MRI, 12 proteins were associated with lower entorhinal thickness (1 FDR, 11 nominal; Table S3; Figure 3D), adjusting for age, sex, and ethnicity. The strongest association was observed for pTau205 (Figure 3E), an emerging marker of tau neurofibrillary tangle pathology.^33,34^ Other notable nominally significant proteins included BDpTau231 and proteins implicated in aging and inflammation, including CCL11, SLIT2, and SFTPD. To assess the regional specificity of these findings, we also examined cortical thickness in the precentral gyrus, a brain region typically spared in aging and early AD.^32^ No proteins survived FDR correction, although 13 showed nominal associations (6 positive and 7 negative, Table S3; Supplementary Figure S1). Notably, no pTau species associated with precentral gyrus thickness. NGF was the only protein nominally associated with thinner cortex in both regions.

### 2.5. Proteins associated with cerebrovascular disease markers

We next examined associations between circulating proteins and two established MRI markers of small vessel disease: white matter hyperintensity (WMH) volume and volume of MRI-visible perivascular spaces in the basal ganglia (BG-PVS) quantified using specialized deep learning models. Models were adjusted for age, sex, ethnicity, and intracranial volume. A total of 27 proteins were positively associated with WMH volume (1 FDR, 26 nominal; Table S3; Figure 3F), with inflammatory proteins prominently represented. TNF showed the strongest association and was the only protein to survive FDR correction (Figure 3G). Other nominally significant inflammatory proteins included CXCL13, IL6R, LGALS3, and ICAM1. By contrast, higher levels of synaptic protein NPTXR were nominally associated with lower WMH burden.

No proteins were associated with BG-PVS at the FDR threshold, although 7 proteins showed nominally significant positive associations (Table S3; Figure 3H). Pro-inflammatory cytokine CXCL10 exhibited the strongest association (Figure 3I), with additional associations involving proteins implicated in inflammation (S100A12, PAFAH1B3) and metabolic or oxidative stress-related processes (HMOX1, HBA1, PRDX6, FAM3B).

### 2.6. Convergent proteomic associations across cognitive and brain measures

To identify proteins associated with multiple aspects of brain aging, we compared associations across all four cognitive and brain MRI markers (Figure 3J). A heatmap of the top 30 proteins, ranked by the number of nominally significant associations and effect sizes, revealed several proteins associated with multiple outcomes. Notably, higher pTau205 was associated with poorer cognition, reduced entorhinal thickness, and greater WMH burden. Higher PAFAH1B3 (involved in synaptic signaling and neurodevelopmental processes) was associated with poorer cognition, greater WMH burden, and greater PVS volume, while higher PRDX6 was associated with poorer cognition and greater PVS volume. Higher NPTX2 was associated with poorer cognition and reduced entorhinal thickness. An exploratory network analysis based on pairwise protein correlations at a threshold of r = 0.35 identified three broad clusters (Extended data Figure 2). One cluster was enriched for AD proteins (including multiple pTau species); a second for predominantly cytokine and chemokine cluster, and a third heterogeneous cluster including inflammatory and cellular stress-related proteins.

Finally, we examined whether plasma proteins explained additional variance in cognition beyond demographic, neuroimaging, and genetic factors. Among the 58 nominally associated with cognition, the 25 proteins with the greatest incremental explanatory power individually explained an additional 2.0–3.6% of variance (Table S4, Extended data Figure 3). When included simultaneously, these 25 proteins collectively explained an additional 14.3% of variance compared with 0.9% for brain MRI markers, 1.0% for *APOE* ε*4* carriage, 25.9% for demographic factors. These exploratory findings suggest that circulating proteins may capture aspects of cognitive variation not accounted for by conventional demographic and neuroimaging measures.

### 2.6. Plasma protein associations with cognition and brain outcomes are supported in an independent cohort

To evaluate the reproducibility of proteins identified in CAMERA, we analyzed data from 1,381 participants in the Alzheimer’s Disease Neuroimaging Initiative (ADNI; mean [*SD*] age = 72.7 [7.24], 726 [52.6%] male; 658 [47.7%] mild cognitive impairment; 213 [15.4%] dementia; 1,311 [94.9%] White; Table S5; Figure 4A). Baseline plasma samples were assayed for 131 proteins on the NULISAseq CNS 120 panel. After quality control (see Methods), 116 proteins and two protein ratios (pTau217/Aβ42; pTau217/MAPT [pTau217%] overlapped with those measured in CAMERA. We examined longitudinal change in two cognitive outcomes: the modified PACC (mPACC)^31^ and CDR sum of boxes (CDR-SB) over a mean follow-up of 4.8 (3.5) years. We also examined two cross-sectional neuroimaging outcomes: cortical thickness and WMH burden. Given the higher prevalence of cognitive impairment in ADNI, we focused on an AD signature region reflecting both early and more advanced AD-related neurodegeneration. Entorhinal cortical thickness was examined as a secondary outcome.

**Figure 4.**
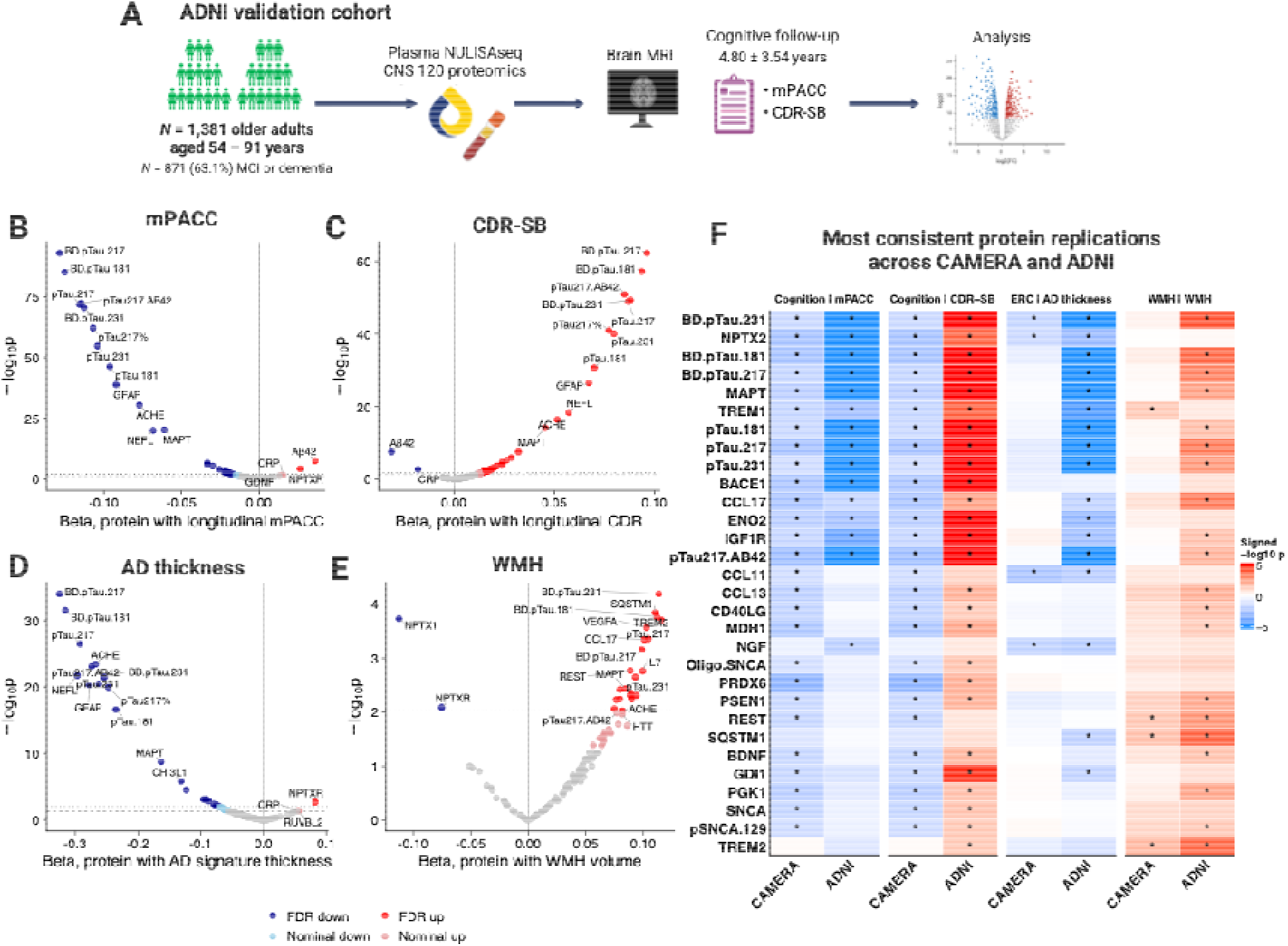
External validation of plasma proteomic associations with cognitive decline and brain MRI measures in ADNI. **A**. Overview of the ADNI validation cohort, comprising 1,381 participants who underwent plasma profiling using the NULISAseq CNS 120 panel, brain MRI, and longitudinal cognitive and clinical assessments. **B.** Associations of baseline plasma proteins with longitudinal change in the modified Preclinical Alzheimer’s Cognitive Composite (mPACC). Higher levels of 36 proteins were associated with faster cognitive decline (27 FDR, 9 nominal), whereas four proteins were associated with slower decline (2 FDR, 2 nominal). **C.** Associations of baseline plasma proteins with longitudinal change in Clinical Dementia Rating Sum of Boxes (CDR-SB). Higher levels of 59 proteins were associated with faster clinical progression (42 FDR, 17 nominal), whereas two proteins were associated with slower progression (both FDR-significant). **D.** Associations of plasma proteins with cortical thickness in Alzheimer’s disease-signature regions. Higher levels of 35 proteins were associated with lower cortical thickness (26 FDR, 9 nominal), whereas 39 were associated with greater cortical thickness (22 FDR, 17 nominal). **E.** Associations of plasma proteins with log-transformed white matter hyperintensity (WMH) volume. Higher levels of 39 proteins were associated with greater WMH volume (22 FDR, 17 nominal), whereas two proteins were associated with lower WMH volume (both FDR-significant). **F.** Heatmap of the 30 proteins showing the strongest cross-cohort concordance in associations with cognitive and MRI outcomes in CAMERA and ADNI, ranked by the number of concordant nominally significant associations and signed −log10(P) values. **Statistical analyses:** Longitudinal associations (B,C) were estimated using linear mixed-effects models with participant-specific random intercepts and slopes, adjusted for baseline age, sex, education, and their interactions with time. Cross-sectional MRI associations (D,E) were estimated using linear models adjusted for age, sex, imaging site, and time between blood collection and MRI, with additional adjustment for intracranial volume in WMH analyses. Asterisks indicate nominal significance: * *p* < .05, ** *p* < .01, *** *p* < .001.

For longitudinal cognitive outcomes, in linear mixed-effects models adjusted for age, sex, and education, 36 proteins (27 FDR, 9 nominal) were associated with faster mPACC decline, including several brain-derived pTau forms: BDpTau217, BDpTau181, and BDpTau231 (Table S6; Figure 4B). Conversely, four proteins (2 FDR, 2 nominal) were associated with slower mPACC decline, including Aβ42 and NPTXR. A similar pattern emerged for CDR-SB, with 59 proteins (42 FDR, 17 nominal) associated with faster progression (Table S6; Figure 4C) including several brain-derived pTau isoforms. By contrast, higher Aβ42 and CRP, were associated with slower CDR-SB progression (both FDR-significant).

For cross-sectional neuroimaging measures, linear models adjusted for age, sex, imaging site, and time between blood collection and MRI identified 35 proteins (26 FDR, 9 nominal) associated with reduced AD-signature cortical thickness, including pTau isoforms, GFAP, NEFL, ACHE, MAPT, and CHI3L1. In contrast, 3 proteins (1 FDR, 2 nominal) were associated with greater cortical thickness, with NPTXR surviving FDR correction (Table S6; Figure 4D). Analyses of entorhinal cortical thickness yielded similar findings (Table S6, Supplementary Figure S2). Thirty-nine proteins (22 FDR, 17 nominal) were associated with greater WMH volume (log-transformed and adjusted for intracranial volume), including BDpTau231, TREM2, VEGFA, and SQSTM1 (Table S6; Figure 4E). By contrast, 2 proteins, NPTX1 and NPTXR (both FDR), were associated with lower WMH burden.

To assess the consistency of protein associations across cohorts, we compared protein effect estimates for cognitive and neuroimaging outcomes in CAMERA and ADNI. Correlation analyses revealed effect size concordance (r = 0.21 – 0.44, Extended data Figure 4). A heatmap highlighting the 30 proteins with the most consistent associations across outcomes and cohorts (Figure 4F) identified several recurring proteins, including BDpTau231 (Extended data Figure 5), NPTX2 (Extended data Figure 6), BDpTau181, BDpTau217, BACE1, TREM1, ENO2, PRDX6, CCL17, and IGF1R.

### 2.7. Cardiometabolic risk factors share proteomic signatures with cognitive and brain aging

Given the established contribution of cardiometabolic risk factors to AD and related dementias,^35,36^ their influence on the circulating proteome,^27,37,38^ and their potential contribution to ethnic group differences in dementia risk,^39,40^ we examined whether cardiometabolic risk and cognitive and brain aging share common proteomic signatures in CAMERA. Identifying these shared signatures may provide insight into the molecular pathways linking cardiometabolic health to cognitive and brain aging. We examined six cardiometabolic factors that together reflect adiposity (i.e., BMI), vascular health (i.e., systolic blood pressure), lipid profiles (i.e., triglycerides, HDL and LDL cholesterol), and glycemic regulation (i.e., HbA1c). Separate linear models adjusted for age, sex, and ethnicity examined associations between each cardiometabolic measure and circulating protein levels.

Cardiometabolic measures showed widespread associations with the circulating proteome, particularly those involved in inflammation and metabolism (Table S7). BMI was associated with 67 proteins (50 FDR, 17 nominal), including positive associations with inflammatory proteins (TNFSF14, CRP, and CCL22), and negative associations with AD biomarkers, including BACE1 and BDpTau217% (Figure 5A). HbA1c was associated with 69 proteins (20 FDR, 49 nominal), including positive associations with the aging marker GDF15 and metabolic proteins NPY and MDH1 (Figure 5B). HDL cholesterol was associated with 65 proteins (40 FDR, 25 nominal). Lower levels of HDL were associated with higher levels of lipid-related proteins such as APOA2 and LDLR and higher levels of inflammatory markers TNFSF14, TNF, CXCL13 (Figure 5C). Triglycerides showed the most extensive associations (88 total, 66 FDR, 22 nominal), including positive associations with LDLR, APOE and several inflammatory proteins, and a negative association with HDL component protein APOA1 (Figure 5D). Associations with systolic blood pressure were comparatively weaker, with no proteins surviving FDR correction, although 42 proteins showed nominal associations including immune targets S100A12, CCL22, CRP and CHI3L1 (Figure 5E). Finally, LDL cholesterol was associated with 39 proteins (8 FDR, 31 nominal), with higher LDL associated with higher levels of APOE, cell adhesion protein ITGAV, and inflammatory protein TNFSF14. Counterintuitively, lower LDL was associated with higher levels of GDF15, BDpTau181, and BDpTau231 (Figure 5F).

**Figure 5.**
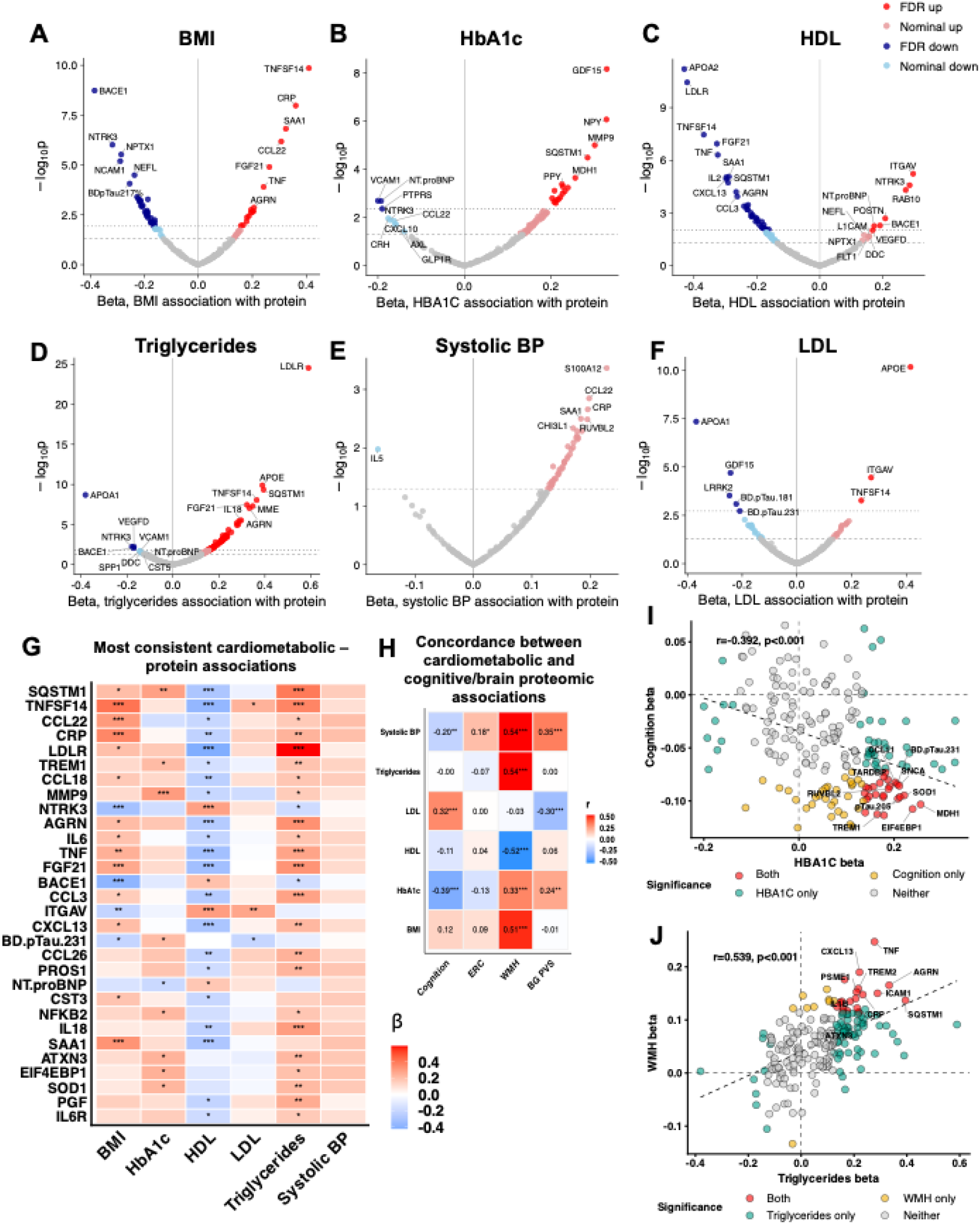
Cardiometabolic factors are associated with plasma proteomic signatures of cognitive and brain aging. **A-E**. Volcano plots depicting results of separate linear models testing associations of cardiometabolic risk factors with proteins, adjusted for age, sex, and ethnicity: **A.** BMI was associated with 67 proteins (50 FDR, 17 nominal); **B.** HbA1c was associated with 69 proteins (20 FDR, 49 nominal); **C.** HDL was associated with 65 proteins (40 FDR, 25 nominal); **D.** Triglycerides were associated with 88 proteins (66 FDR, 22 nominal); **E.** Systolic blood pressure was not associated with any proteins at the FDR threshold, but with 42 proteins nominally. **F.** LDL was associated with 39 proteins (8 FDR, 31 nominal). **G.** Heatmap depicting top 30 proteins that most consistently associated with cardiometabolic factors, ranked by number of FDR-significant associations and effect size. **H.** Heatmap depicting Pearson correlations between protein effect sizes for cognitive/brain outcomes (x-axis) and cardiometabolic factors (y-axis). **I.** Effect size scatter plots for hbA1c associations with proteins (x-axis) and protein associations with cognition (y-axis). **J.** Effect size scatter plots for triglycerides levels associations with proteins (x-axis) and protein associations with white matter hyperintensity (WMH) volume (y-axis). The asterisks reflect FDR-corrected p-values. * *p* < .05, ** *p* < .01, *** *p* < .001.

To identify shared proteomic patterns across cardiometabolic measures, we examined the 30 proteins showing the most consistent associations (Figure 5G). These proteins predominantly involved immune and inflammatory processes, including CRP, TNF, TNFSF14, CCL22, CCL18, MMP9, and IL6. Many showed positive associations with BMI, HbA1c, and triglycerides, and negative associations with HDL cholesterol, consistent with a common inflammatory signature of adverse cardiometabolic health. Several brain-relevant proteins, including TREM1, NTRK3, BACE1, and SAA1, were among the top 30, suggesting shared biological pathways underlying cardiometabolic and brain health.

We next examined whether proteins associated with cardiometabolic risk overlapped with those associated with cognition and MRI measures of brain aging in CAMERA. To assess this overlap, we correlated protein effect estimates from the cardiometabolic and cognitive/neuroimaging models (Figure 5H). Proteins associated with greater WMH burden tended to show associations with cardiometabolic measures in directions consistent with greater cardiometabolic risk, except LDL cholesterol. Similarly, proteins associated with greater BG-PVS volume showed concordant positive associations with higher HbA1c and systolic BP, whereas proteins associated with better cognitive performance tended to be associated with lower HbA1c and lower systolic BP. However, not all patterns followed expected risk directions. Protein effect estimates for entorhinal cortical thickness were positively correlated with those for systolic blood pressure, while estimates for LDL cholesterol were positively correlated with better cognition and negatively correlated with BG-PVS volume.

At the individual protein level, several proteins showed overlapping associations with cardiometabolic and brain aging outcomes. Higher pTau205, TREM1, CCL11, and BDpTau231 were associated with both higher HbA1c and poorer cognition (Figure 5I), whereas higher TNF, TREM2, AGRN, and CRP were associated with both higher triglycerides and greater WMH burden (Figure 5J). Together, these findings highlight shared protein signatures of cardiometabolic risk and adverse cognitive and neuroimaging outcomes, with inflammatory pathways prominently represented.

### 2.8. Sensitivity analyses

In CAMERA, associations between plasma proteins with cognitive, brain and cardiometabolic measures remained largely unchanged after additional adjustment for *APOE* ε*4* carrier status (Table S8; Supplementary Figures S3 and S4). Associations with cardiometabolic factors were also largely unchanged after adjustment for relevant medication use (i.e., lipid-modifying medications, glucose-lowering medications, blood pressure-lowering medications; Table S9; Supplementary Figure S5) and BMI (Table S10; Supplementary Figure S6). In ADNI, excluding participants with dementia at baseline (*N*=213) yielded results largely consistent with the primary analyses for cognitive and neuroimaging outcomes (Table S11, Supplementary Figure S7).

## 3. DISCUSSION

In an ethnically diverse cohort with substantial representation of Chinese and South Asian adults, populations historically underrepresented in aging and AD biomarker research, we used the expanded NULISAseq 220 panel to characterize circulating proteomic signatures of cognitive performance, cortical thickness, cerebral small vessel disease, and cardiometabolic health. We identified established and emerging markers of AD pathology, synaptic function, inflammation, and oxidative stress, including brain-derived p-tau species, pTau205 and pTau212,^17–21^ plus targets such as BACE1, NPTX2, TREM1, PAFAH1B3, and PRDX6. Several proteins showed associations across multiple cognitive and neuroimaging outcomes, with findings also supported in an independent cohort with substantially different demographic and clinical characteristics. We further identified considerable overlap between proteomic signatures of poorer cognitive performance, greater cerebral small vessel disease burden and cardiometabolic risk, particularly involving inflammatory proteins Together, these findings expand our understanding of circulating proteomic signatures of cognitive and brain aging to ethnically diverse populations and identify biological processes shared with cardiometabolic health.

Using the expanded NULISAseq Neuro 220 panel, we identified widespread associations of circulating proteins with age, sex, and *APOE* ε4 status, extending previously reported and emerging proteomic patterns to a broader range of brain-relevant proteins in an ethnically diverse cohort.^41–43^ Older age was associated with higher levels of proteins implicated in neurodegenerative disease and neurodegeneration (pTau, GFAP, NEFL, NEFH), inflammation (TREM1, TREM2, CHI3L1), cardiac stress (NTproBNP), and systemic aging (GDF15).^2^ We also observed sex differences in several brain-relevant proteins, including the synaptic proteins NPTX1 and NPTX2, which were higher in women and men, respectively. A similar pattern was reported by Trelle and colleagues in cohort of cognitively unimpaired older adults.^13^ *APOE* ε4 carriers had higher levels of several AD-related proteins, including brain derived pTau species, the amyloid processing protein BACE1, and the synaptic protein GAP43. Exploratory analyses suggested that the magnitude of *APOE* ε4-related differences in some pTau species may vary across ethnic groups, with generally stronger associations among non-Hispanic White than among Chinese and South Asian participants. In contrast, *APOE* ε4 associations with BACE1 and GAP43 appeared more similar across ethnic groups. These findings are consistent with a recent study reporting smaller *APOE* ε4-associated increases in plasma pTau217% among Asian compared to White older adults.^44^ Although limited by small subgroup sizes and exploratory interaction analyses, our findings raise the possibility that *APOE* ε4-related proteomic signatures differ across populations. Larger studies are needed to establish whether these differences are reproducible and relevant to the interpretation of blood-based AD biomarkers.

Several circulating proteins were associated with poorer cognitive performance, including established AD biomarkers as well as proteins involved in synaptic function and oxidative stress. Associations involving pTau217 and brain-derived pTau species are consistent with previous studies linking circulating tau biomarkers to cognitive impairment and decline.^45,46^ Beyond these AD markers, we identified associations involving NPTX2,^47^ STX4,^48^ and PRDX6.^49^ Notably, higher plasma NPTX2 was associated with poorer cognition, contrasting with previous studies linking lower CSF NPTX2 to faster cognitive decline and progression to MCI.^50,51^ Since CSF and plasma levels of NPTX2 do not correlate,^52^ associations of peripheral NPTX2 with cognitive outcomes may partly reflect processes linked to non-central nervous system expression of NPTX2 in tissues such as the pancreas, adrenal glands, and testes (in men). Higher plasma STX4, a protein involved in vesicular trafficking, was also associated with poorer cognition. PRDX6, an antioxidant enzyme involved in cellular membrane homeostasis, has been implicated in AD dementia,^53^ with increased expression reported in experimental models of AD^49^ and in hippocampal tissue from individuals with AD.^54,55^ Our findings extend these observations to plasma, identifying PRDX6 as a correlate of cognitive performance in a predominantly cognitively unimpaired, ethnically diverse cohort.

Among the neuroimaging outcomes, plasma pTau205 showed the strongest association with lower entorhinal cortical thickness and was the only protein to survive FDR correction. The entorhinal cortex is among the earliest regions affected by AD,^56,57^ but is also vulnerable to age-related tauopathy.^58^ CSF studies suggest that pTau205 becomes abnormal later in the AD continuum than other phosphorylated tau species and may more closely reflect neurofibrillary tangle pathology.^21,33,34^ Our findings extend this work to plasma, identifying an association between pTau205 and entorhinal thinning in a predominantly cognitively unimpaired cohort, although whether this reflects AD-related or age-related tau pathology remains unclear.

In contrast, proteomic associations with cerebral small vessel disease were characterized predominantly by inflammatory proteins. TNF showed the strongest association with WMH burden and was the only protein to survive FDR correction. Other nominal associations included proteins involved in immune signaling (CXCL13 and IL6R), vascular inflammation (ICAM1), and inflammatory and tissue-remodeling processes (LGALS3).^59^ These findings are consistent with evidence implicating inflammatory signaling, endothelial dysfunction, and immune activation in cerebral small vessel disease.^60,61^ Associations with MRI-visible basal ganglia PVS were weaker, with no proteins surviving correction for multiple comparisons, although nominal associations involving inflammatory and oxidative stress-related proteins were observed. These findings may reflect processes involved in perivascular and vascular homeostasis,^62^ although the evidence remains preliminary. The different proteomic patterns associated with entorhinal thickness, WMH, and PVS suggest that these measures capture partly distinct aspects of structural brain aging. However, the limited number of associations surviving multiple-comparison correction warrants caution.

An important finding was that several proteins were associated with more than one cognitive or neuroimaging outcome, including pTau205, NPTX2, PRDX6, and PAFAH1B3, a protein involved in neurodevelopmental and cellular signaling.^63^ These findings suggest that some circulating proteins may reflect biological processes shared across different aspects of brain aging. We also explored whether plasma proteins provided information about cognitive performance beyond demographic characteristics, *APOE* ε4 status, and neuroimaging measures. In exploratory analyses, 25 selected proteins collectively explained an additional 14.3% of the variance in cognition. However, because these proteins were selected and evaluated in the same cohort, the estimate may be inflated and requires independent validation.

Analyses in ADNI using the NULISAseq CNS 120 panel provided independent support for several of the protein associations identified in CAMERA, despite substantial differences in ethnic composition, cognitive status, and outcome measures. Consistent patterns were observed for established AD-related proteins, including brain-derived pTau species and pTau217, as well as proteins involved in synaptic function, inflammation, and cellular processes, including NPTX2, TREM1, CCL17, PRDX6, and IGF1R. Notably, higher plasma NPTX2 was associated with poorer cognitive performance in CAMERA and worse longitudinal cognitive outcomes in ADNI, providing further support for the direction of the plasma association despite its contrast with previous CSF findings. Although correlations between protein effect estimates across cohorts were moderate, differences in cohort characteristics and outcome measures may have contributed to this variability. The consistency of several individual associations is nevertheless notable given these differences. Comparisons were limited to proteins shared between the CNS 120 and Neuro 220 panels, leaving additional targets, particularly pTau205, without independent validation.

Beyond cognitive and neuroimaging measures, we identified substantial overlap between signatures of cardiometabolic risk and brain aging. Proteins associated with greater WMH burden generally showed associations with poorer cardiometabolic health whereas those associated with worse cognition tended to be associated with higher HbA1c and systolic blood pressure. At the individual protein level, higher pTau205, TREM1, CCL11, and BDpTau231 were associated with both higher HbA1c and poorer cognitive performance, whereas higher TNF, TREM2, AGRN, and CRP were associated with higher triglycerides and greater WMH burden. The prominence of inflammatory proteins across these associations is consistent with growing evidence linking cardiometabolic dysfunction and inflammation to neurodegenerative and cerebrovascular disease.^64,65^ Higher BMI was associated with lower levels of plasma BACE1 and BDpTau217%. The latter finding is consistent with previous reports of inverse associations between later-life BMI and AD pathology or progression.^66,67^ Such associations may reflect reverse causation, whereby weight loss accompanies preclinical neurodegeneration, rather than a protective effect of higher adiposity.^68^

This study has several strengths, including the use of the expanded NULISAseq Neuro 220 panel in a deeply phenotyped, ethnically diverse cohort with detailed neuropsychological, neuroimaging, and cardiometabolic assessments. Importantly, the substantial representation of Chinese and South Asian participants extends plasma proteomic research to populations historically underrepresented in dementia biomarker studies. Independent analyses in ADNI further allowed us to assess the consistency of several protein associations in a cohort with different demographic and clinical characteristics.

Several limitations warrant consideration. First, although CAMERA is an ongoing longitudinal study, the present analyses were restricted to baseline data, precluding assessment of whether the identified proteomic signatures predict subsequent cognitive decline, atrophy, or cerebral small vessel disease progression. Second, although CAMERA included substantial representation of Chinese and South Asian participants, the sample size limited statistical power to robustly investigate ethnic differences in proteomic associations. Third, without direct measures of cerebral amyloid and tau pathology, we cannot determine whether associations involving AD-related proteins reflect underlying AD pathology. Fourth, circulating proteins may originate from multiple tissue sources and their plasma concentrations do not necessarily reflect changes within the brain. Finally, external validation was restricted to 118 proteins and ratios shared between CAMERA and ADNI, leaving several novel targets without independent validation. Longitudinal follow-up in CAMERA and replication in larger, ethnically diverse cohorts will be important for establishing the reproducibility and clinical relevance of these findings.

In conclusion, application of the expanded NULISAseq Neuro 220 platform identified novel circulating proteomic signatures of cognitive and brain aging in an ethnically diverse cohort with substantial representation of Chinese and South Asian adults. Beyond established AD biomarkers, we identified proteins associated with cognitive performance and structural brain changes, spanning synaptic function, oxidative stress, and inflammation, with several associations independently supported in ADNI. The substantial overlap between brain-related and cardiometabolic proteomic signatures further highlights shared inflammatory and metabolic processes associated with brain aging. Together, these findings expand our understanding of the circulating proteome in brain aging and underscore the potential of multiplex plasma proteomics to advance biomarker discovery and more inclusive approaches to dementia risk assessment.

## Data Availability

All data produced in the present study are available upon reasonable request to the authors.

## Funding/acknowledgements

This work was supported by funding from the Canadian Institutes of Health Research (173253; JSR and MG), the Alzheimer Society of Canada New Investigator Award (22-25, JSR), and the Alzheimer’s Association (AARG-23-1144933; JSR). Additional support comes from the Ontario Ministry of Colleges, Universities, Research Excellence and Security (Early Researcher Award, ER24-18-113; JSR), the Larry L. Hillblom Foundation (2024-A-001-CTR; PI: KBC), New Vision Research (CCAD 2024-001-1; PI: RS), an award from the American Academy of Neurology (PI: RS). BW is supported by the Queen Elizabeth II Graduate Scholarship in Science & Technology. MWA, TM, and KV are supported by the Canada Graduate Scholarships program. MWA and RY are supported by the Alzheimer’s Society of Canada. MG is supported by the Canada Research Chairs program (CRC-2021-00374). Computational resources for this work were supported by funding from the Canada Foundation for Innovation (40206, 45908, 43890, 43910) and the Ontario Research Fund (MG).

## Code Availability

Algorithms used for data analyses are accessible in publicly available, existing R and Python packages and code repositories. Freesurfer v7, ICVMapp3r, and segcsvd are all open-source/open-weight neuroimaging tools, available at the following links: Freesurfer: https://surfer.nmr.mgh.harvard.edu/ ICVMapp3r: https://github.com/AICONSlab/iCVMapp3r segcsvd: https://github.com/AICONSlab/segcsvd

## 4. METHODS

### 4.1 The CAMERA study

The Canadian Multi-Ethnic Research on Aging (CAMERA) study is a Toronto-based cohort comprising adults aged 55-85 who self-identify as South Asian, Chinese, or non-Hispanic White. The study design eligibility criteria have been described previously.^23^ CAMERA recruited community-dwelling adults with the aim of enrolling cognitively unimpaired individuals (Clinical Dementia Rating [CDR] global score of 0); however, participants subsequently found to have a CDR global score of 0.5 following enrollment were retained. Exclusion criteria included major neurological or psychiatric disorders, unstable medical conditions, and insufficient English proficiency to provide informed consent and complete study assessments. Participants underwent blood collection, neuropsychological and clinical assessments, and a brain magnetic resonance imaging (MRI) scan. The study was approved by the Research Ethics Board of Sunnybrook Health Sciences Centre, and all participants provided written informed consent.

### 4.2 Plasma NULISAseq proteomics

Plasma was collected following an overnight fast into ethylenediaminetetraacetic acid (EDTA) tubes and stored at -80°C until analyses. Plasma proteomics were quantified using the NULISAseq Neuro 220 Panel (Alamar Biosciences, Fremont, California, USA), which measures 220 proteins relevant to brain health and neurological disease.^13–16^ Samples were thawed and centrifuged at 2,200g for 10 minutes. Using the automated Alamar Argo HT platform, immunocomplexes were generated and labeled with sample- and target-specific DNA barcodes, pooled into a single library and quantified by next-generation DNA sequencing. Samples were randomized across plates, stratified by ethnicity, and analyzed in singlicate, according to previously established NULISA procedures.^15,16^

All quality control (QC) metrics met predefined thresholds, including median intra-plate internal control coefficient of variation (CV) and median inter-plate control CV of 5.94% (threshold of 10%) and 6.99% (threshold of 15%), respectively. In CAMERA, 11 proteins (Aβ38, CALB2, CGRP, FCGR2B, GSDMD, PARP1, pLRRK2.S1292, PTN, SNCB, UCHL1, and YWHAZ) with median detectability below 50% were excluded, consistent with previous NULISAseq studies.^16,69^ Seven proteins associated with rare genetic disorders were also excluded (pQ.HTT, mHTT.exon1, pPRKN.S65, pQ.ATXN3, pRAB10.T73, pRAB12.S106, pRAB29.T71).

APOE4 and CHIT1 were excluded from continuous protein association analyses due to their bimodal distributions. However, APOE4 was used to infer *APOE* ε4 allele carrier status. To minimize the influence of extreme values, protein measurements were winsorized at three times the interquartile range. We used the weighted gene co-expression network analysis (WGCNA) sample network connectivity outlier algorithm to identify sample outliers (defined as >3 *SD* below mean sample connectivity), resulting in the removal of two samples from the analytic proteomics dataset.^70,71^

Following these exclusions, analyses included 200 individual NULISAseq proteins and three protein ratios reflecting AD pathology (pTau217/AB42, pTau217%, and BDpTau217%), calculated as the difference between log-transformed NULISA Protein Quantification (NPQ) values, as previously described.^20^ To facilitate interpretation and effect size comparison, all 203 proteomic measures were z-transformed prior to analysis.

### 4.3 *APOE* ε4 carrier status

In CAMERA, APOE4 protein levels were used to infer APOE ε4 carrier status, with participants classified as carriers (APOE4 > 4 NPQ) or noncarriers (APOE4 ≤ 4 NPQ; Supplementary Figure S8).

### 4.4 Cognitive assessments

CAMERA participants completed a battery of cognitive tests assessing attention, executive function, processing speed, episodic memory, language, and visuospatial abilities. Cognitive performance was quantified using a composite informed by the Preclinical Alzheimer Cognitive Composite (PACC),^31^ emphasizing measures sensitive to early AD-related cognitive changes^72,73^ and incorporating predominantly nonverbal tasks to minimize language-related influences across ethnic groups. The composite included the Free and Cued Selective Reminding Test total score (FCSRT96; sum of free and cued recall across three learning trials),^74^ the Doors Subtest total score (Part A and B) from the Doors and People Test,^75^ category fluency (Animals),^76^ the Digit Symbol Coding subtest (WAIS-III),^77^ and the Mnemonic Similarity Test Set C (lure discrimination index, LDI).^78^ Scores on each test were z-transformed and averaged to compute the composite, with higher scores reflecting better performance.

Global cognitive and functional status was assessed using the CDR based on interviews with participants and their study partners. All assessments were administered by trained research staff under the supervision of a clinical neuropsychologist.

### 4.5 Brain MRI

*N* = 233 CAMERA participants underwent brain MRI on a 3T Siemens MAGNETOM Prisma scanner with a 64-channel head coil. Axial 3D T1-weighted (TR=2400 ms, TE=2.28 ms) magnetization-prepared rapid gradient echo (MPRAGE) images were acquired at 0.8 mm isotropic resolution with a 256x256x180 mm field of view (FOV). T1-weighted images were skull-stripped using a deep learning tool (ICVMapp3r^79^) and processed with FreeSurfer v7,^80^ including intensity normalization, white and pial tissue segmentation, and cortical parcellation using the Desikan-Killiany atlas.^81^

Sagittal 3D T2-weighted (TR=5000 ms, TE=388 ms) fluid attenuated inversion recovery (FLAIR) images were acquired at 1.0 mm isotropic resolution and with a 256x256x192 mm FOV. FLAIR images were rigidly co-registered to the T1-weighted images followed by automated segmentation of white matter hyperintensity (WMH) and enlarged perivascular space (PVS) using a deep-learning based tool, segcsvd.^82–84^ Basal ganglia (BG) PVS volumes were generated by intersecting the PVS segmentation with a BG mask generated from FreeSurfer subcortical segmentations (caudate, putamen, pallidum, and accumbens nucleus).

We focused on three MRI-derived measures to capturing complementary aspects of brain aging. Mean bilateral entorhinal cortical thickness was selected as a marker of early neurodegeneration, given its vulnerability to age and AD-related atrophy.^56,57^ Mean bilateral precentral gyrus thickness was examined as an AD-spared control region. We also examined two markers of cerebral small vessel disease: WMH volume and BG-PVS volume, both log-transformed to reduce skewness and corrected for intracranial volume in analyses. To facilitate interpretation, all three measures were z-transformed prior to analyses.

### 4.6 Cardiometabolic measures

Cardiometabolic health was assessed using standardized clinical measurements, including body mass index (BMI), calculated from measured height and weight, systolic blood pressure (averaged across three seated readings taken one minute apart using an automated sphygmomanometer), plus serum levels of triglycerides, high-density lipoprotein (HDL) and low-density lipoprotein (LDL) cholesterol, and HbA1c. Medication use was self-reported, and lipid-modifying, glucose-lowering, and antihypertensive medications were included as covariates in sensitivity analyses.

### 4.7 External validation cohort

To externally validate associations between plasma proteins with cognitive and brain outcomes identified in CAMERA, we analyzed NULISAseq data from the Alzheimer’s Disease Neuroimaging Initiative (ADNI; adni.loni.usc.edu). The primary goal of ADNI has been to test whether serial neuroimaging, biological markers, and clinical and neuropsychological assessment can be combined to measure the progression of mild cognitive impairment (MCI) and early Alzheimer’s disease (AD). Given differences in cohort composition and outcome measures, validation focused on the consistency of protein associations with corresponding cognitive and neuroimaging outcomes rather than exact replication of the CAMERA analyses.

Plasma samples from 1,428 ADNI participants were assayed using the NULISAseq CNS 120 panel, comprising 131 protein targets, following standard processing protocols. Consistent with CAMERA, protein concentrations were winsorized at three times the interquartile range. Nine targets were excluded because plate-level detectability was below 50% on more than 20% of plates (SNCB, UCHL1, PTN, YWHAZ, YWHAG, pTDP43409, IL1B, BDMAPT, SNAP25) and APOE4 was excluded because of its bimodal distribution. An additional 14 participants were excluded as sample outliers detected with the WGCNA sample network connectivity outlier algorithm.^70,71^ The resulting dataset included 116 individual proteins, and two derived ratios (pTau217/Aβ42, pTau217%), with 118 proteomic measures overlapping between ADNI and CAMERA. All proteomic measures were z-transformed prior to analysis.

ADNI participants underwent repeated cognitive and clinical assessments every 6-12 months. Longitudinal cognitive performance was assessed using the modified PACC (mPACC),^31,85^ calculated by averaging sample-based z-scores from the Alzheimer Disease Assessment Scale—Cognitive Subscale Delayed Word Recall, Logical Memory Delayed Recall, Mini Mental State Examination (MMSE), and log-transformed Trail-Making Test B Time to Completion, with scores oriented such that higher values indicate better performance. Clinical progression was assessed using longitudinal CDR Sum of Boxes (CDR-SB) scores, which was log-transformed to reduce skewness.

For neuroimaging analyses, we used publicly available processed MRI measures. Given the greater proportion of cognitively impaired participants in ADNI compared with CAMERA (Table S4), the primary neurodegeneration outcome was mean bilateral cortical thickness across AD-vulnerable regions, including the entorhinal, parahippocampal, inferior and middle temporal, and fusiform cortices. Mean bilateral entorhinal thickness, corresponding directly to the primary CAMERA measure, was also examined in secondary analyses.

Total WMH burden was derived from an automated segmentation pipeline. In brief, 3D T1 and FLAIR images were co-registered using an affine transformation, corrected for intensity inhomogeneity, and transformed to a common template. WMH were segmented using a modified Bayesian approach using a likelihood estimation from histogram and threshold-based segmentation, a prior probability mask generated from semi-automated WMH segmentations from a training dataset, and tissue constraints. The final masks were binarized using a 3.5 standard deviation threshold and transformed back into native space.

Given that only cross-sectional FreeSurfer v7 measures were available, we restricted MRI analyses to cross-sectional data, using the first-available brain MRI obtained at or after the time of the NULISA proteomics blood draw, with analyses adjusted for this time difference (0–13.2 years, median = 0.5 years). All cognitive and neuroimaging measures were standardized to facilitate interpretation. The final validation sample included 1,381 participants with available cognitive or MRI data, with a mean (*SD*) cognitive and clinical follow-up of 4.8 (3.5) years.

### 4.8 Statistical analyses

Analyses were conducted in R v4.6.1 and Python v3.11.12. Statistical significance was assessed using a false discovery rate (FDR) threshold of 5%, with multiple comparisons corrected using the Benjamini-Hochberg procedure. Associations meeting nominal (i.e., unadjusted) significance were also reported. Demographic and clinical characteristics were summarized using descriptive statistics and compared using analysis of variance (ANOVA), t-tests, or χ² tests, as appropriate. Correlation analyses and scatterplots were used to assess concordance of effect estimates where specified.

In CAMERA, we first characterized associations of demographic factors with the NULISAseq Neuro 220 proteome. Separate multivariable linear models examined associations of age and sex with individual protein levels, adjusting for ethnicity. We also tested associations of *APOE* ε*4* carrier status (carrier vs. non-carrier) with protein levels, adjusting for age, sex, and ethnicity. Given evidence that associations of *APOE* ε*4* with AD risk and biomarkers may differ across ancestral populations,^30^ we conducted exploratory post hoc stratified and interaction analyses testing whether associations of *APOE* ε*4* carriage and protein levels differed by self-reported ethnicity (South Asian, Chinese, and non-Hispanic White).

We next examined associations of plasma proteins with cognitive performance and MRI-derived measures of brain aging. Separate multivariable linear models were fitted for each protein–outcome pair, adjusting for age, sex, and ethnicity. Cognitive models additionally adjusted for years of education and English as a first language (EFL), whereas volumetric MRI models (i.e., WMH and BG-PVS) additionally adjusted for total intracranial volume. Exploratory variance partitioning analysis examined whether the proteins nominally associated with cognition in main models explained additional variance in cognitive scores beyond demographic variables (i.e., age, sex, years of education, ethnicity, English-as-a-first-language status), *APOE* ε*4* carriage, and neuroimaging measures (i.e., entorhinal thickness, intracranial volume-adjusted WMH and BG-PVS volumes).

To externally validate CAMERA findings, we examined associations of overlapping plasma proteins with corresponding cognitive and MRI outcomes in ADNI. Longitudinal associations with mPACC and CDR-SB were assessed using linear mixed-effects models with participant-specific random intercepts and slopes. Models included protein levels, time, and their interaction, adjusting for baseline age, sex, years of education, and their interactions with time. The protein-by-time interaction was the primary term of interest, reflecting the association between baseline protein levels and longitudinal cognitive or clinical change. Cross-sectional associations with cortical thickness and WMH were assessed using linear models to test associations between protein levels and brain outcomes, adjusting for age, sex, imaging site, time between blood draw and MRI, and intracranial volume (for WMH volume only).

To probe the potential influence of cardiometabolic health on the proteomic signatures of cognitive and brain aging in CAMERA, we also characterized NULISAseq Neuro 220 protein associations with cardiometabolic measures. Separate multiple linear models examined associations of standardized BMI, HbA1c, HDL cholesterol, LDL cholesterol, triglycerides, and systolic blood pressure with individual protein levels, adjusting for age, sex, and ethnicity.

Sensitivity analyses examined the robustness of CAMERA protein associations with cognitive and brain outcomes as well as cardiometabolic factors following additional adjustment for *APOE* ε*4* carrier status. Cardiometabolic models were adjusted for relevant medication use (glucose-lowering medications for HbA1c, lipid-modifying medications for HDL and LDL cholesterol and triglycerides, and antihypertensives for systolic blood pressure) and BMI for all non-BMI predictors. In ADNI, sensitivity analyses re-estimated associations of proteins with cognitive and brain outcomes excluding participants with dementia at baseline.

---

**Extended Data Fig. 1.**
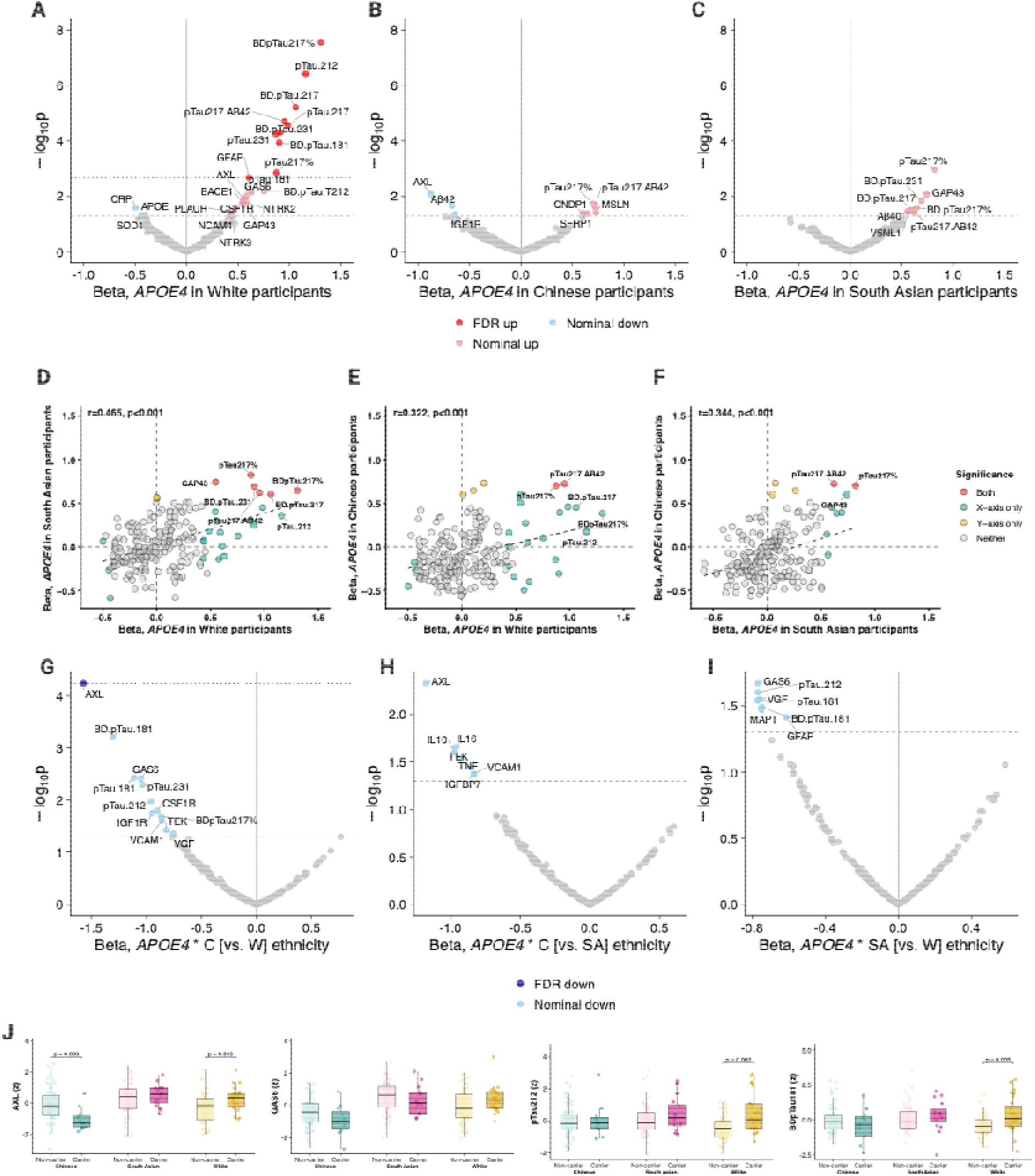
Ethnic differences in APOE ε4-associated plasma proteomic profiles. The volcano plots show associations of *APOE* ε*4* carrier status (vs. non-carrier) with plasma proteins in **A**. non-Hispanic White participants (*N* = 83); **B**. Chinese participants (*N* = 82); and **C**. South Asian (*N* = 74) participants from CAMERA. Associations were estimated using linear models adjusted for age and sex. **D–F**. Scatterplots comparing protein association effect estimates between **D**. non-Hispanic White and South Asian; **E**. non-Hispanic White and Chinese; and **F**. South Asian and Chinese. **G-I**. Volcano plots showing the interactions between ethnicity and *APOE* ε*4* carrier status on protein levels. **J.** Boxplots showing protein levels for top interaction hits by ethnicity and *APOE* ε*4* carrier status. C, Chinese; SA, South Asian; W, White.

**Extended Data Fig. 2.**
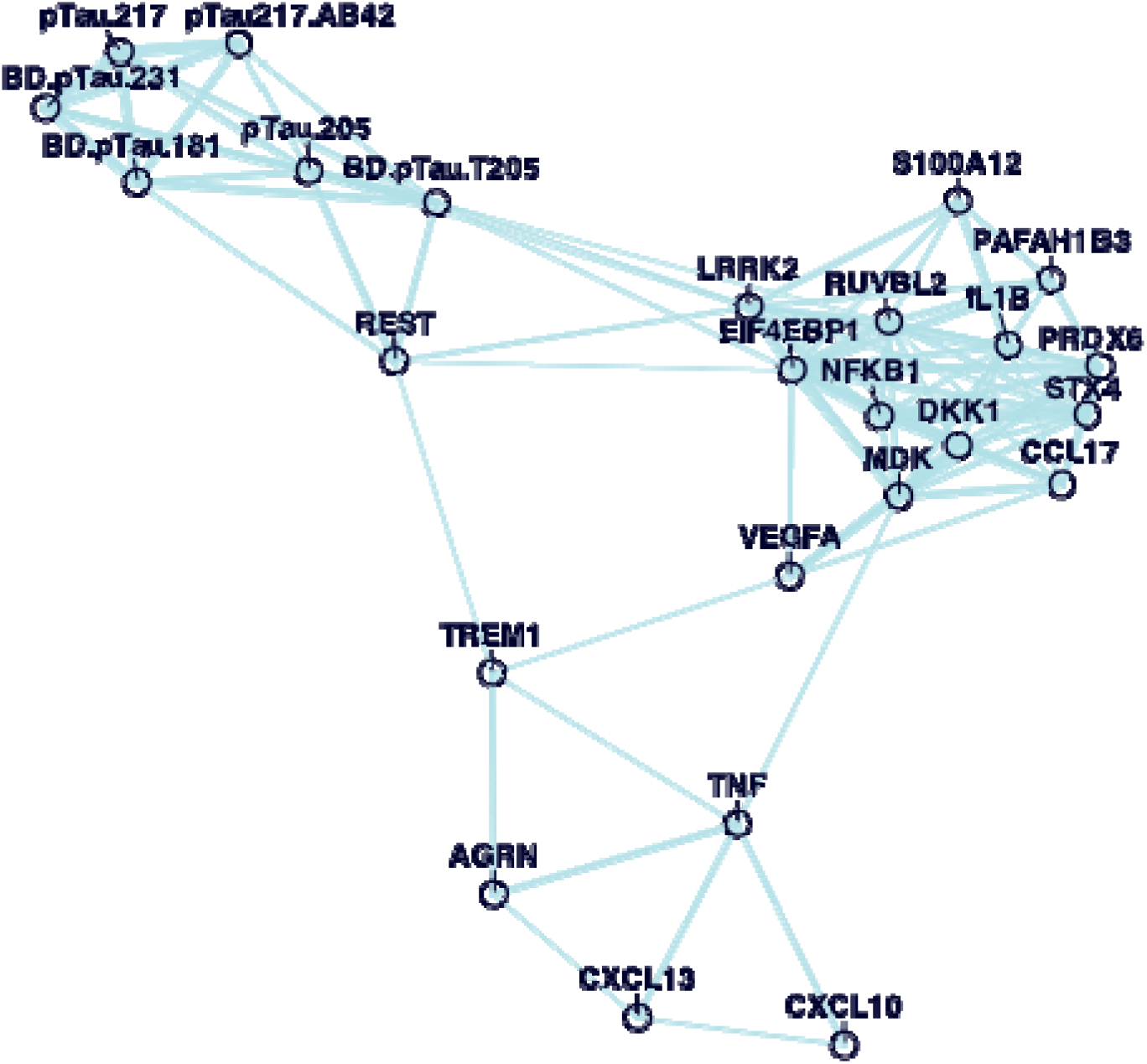
Network plot of top 30 proteins that most consistently associated with cognitive and brain outcomes in CAMERA. A network analysis estimating pairwise correlations at a threshold of r = 0.35 illustrated that proteins associated with cognitive and brain aging cluster into three groups, including a p-tau cluster (e.g., pTau217, BDpTau231, pTau205), a predominantly cytokine/chemokine and immune cluster (e.g., TNF, CXCL13, TREM1), and a more heterogeneous cluster including inflammatory and cellular stress-related biology (e.g., LRRK2, PRDX6, S100A12, among others).

**Extended Data Fig. 3.**
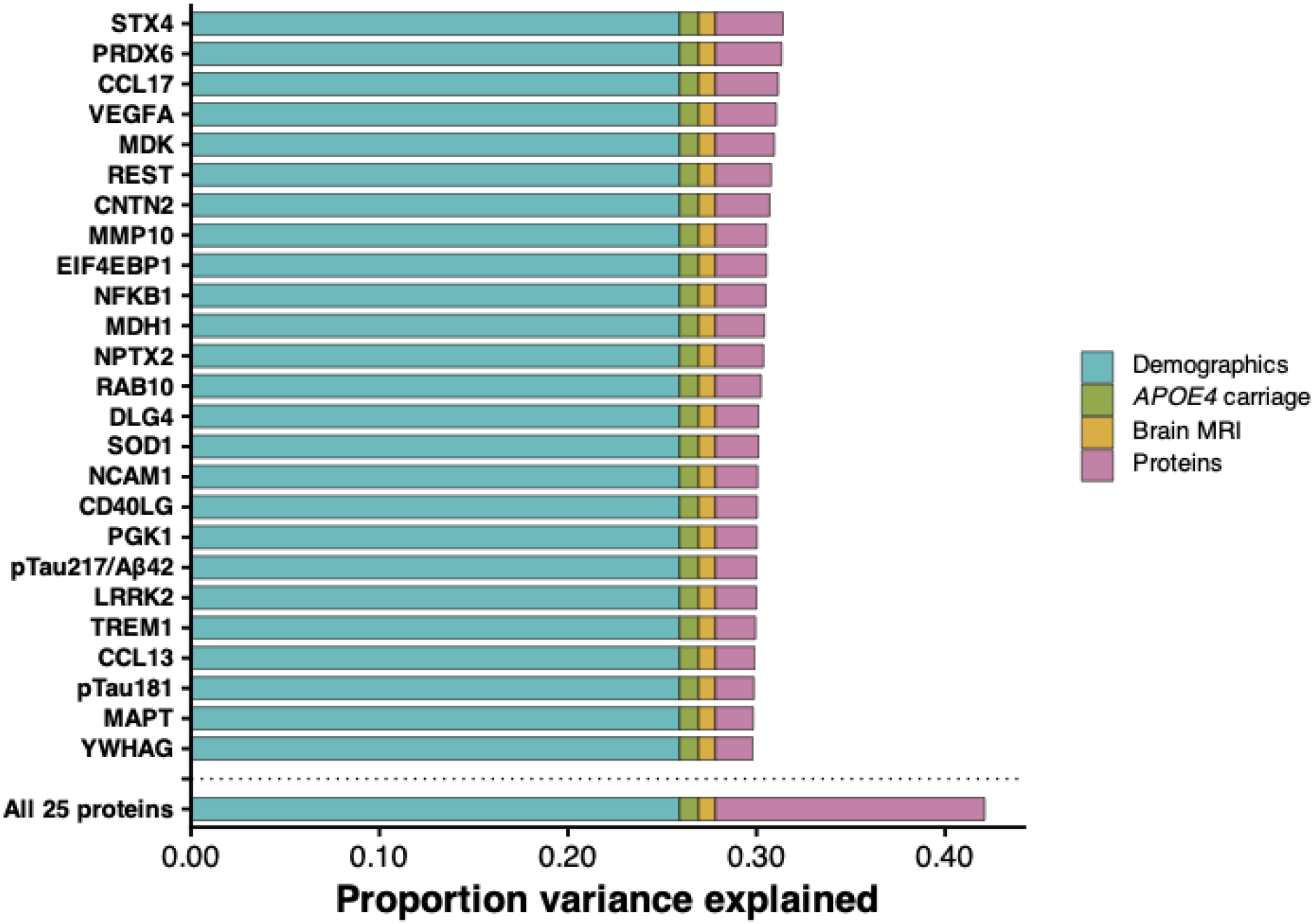
Variance partition plot showing variance in cognitive scores explained by plasma proteins above and beyond demographic factors and brain MRI measures in CAMERA. The plot depicts the top 25 proteins with the greatest incremental explanatory variance in cognitive scores, selected from all proteins nominally associated with cognition in main models. Models include terms for age, sex, years of education, ethnicity, English-as-a-first-language status, *APOE4* carriage, entorhinal thickness, white matter hyperintensity (WMH) volume, basal ganglia perivascular space (BG-PVS) volume, and intracranial volume. Each protein was added individually to the base model to estimate its contribution to explained variance. All 25 proteins were also added simultaneously in a combined model to estimate their total contribution to explained variance.

**Extended Data Fig. 4.**
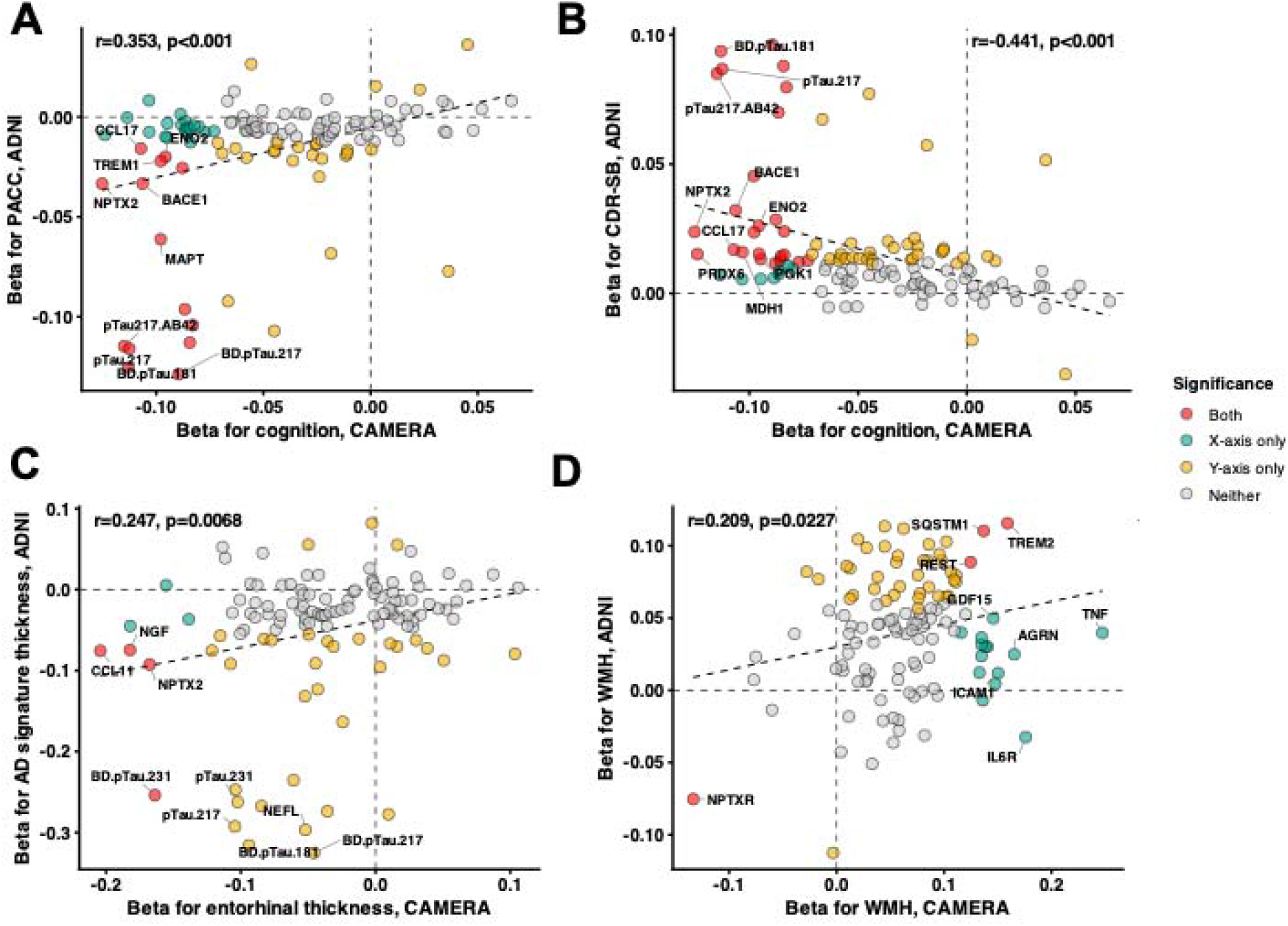
Cross-cohort concordance of plasma proteomic associations with cognitive and MRI outcomes. **A-B.** Scatterplots comparing protein association effect estimates for cognitive performance in CAMERA with **A.** longitudinal changes in modified Preclinical Alzheimer Cognitive Composite (mPACC) and **B.** Clinical Dementia Rating Sum of Boxes (CDR-SB) in ADNI. **C.** Comparison of protein association effect estimates for entorhinal cortical thickness in CAMERA and AD-signature cortical thickness in ADNI. **D.** Comparison of protein association effect estimates for white matter hyperintensity (WMH) volume in CAMERA and ADNI. CAMERA effect estimates are shown on the x-axis and ADNI effect estimates on the y-axis.

**Extended Data Fig. 5.**
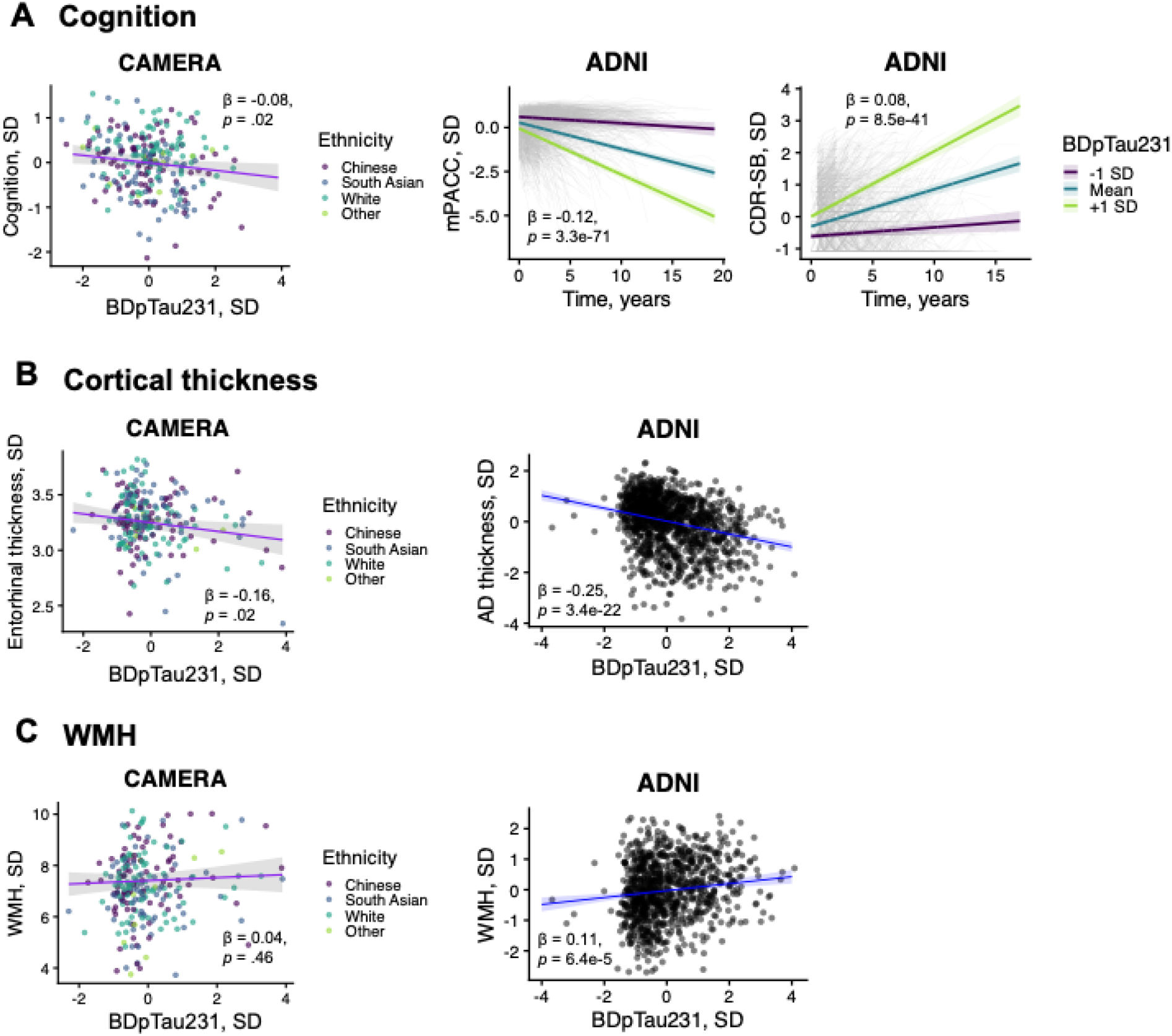
Associations of brain-derived phosphorylated tau 231 with cognitive and MRI outcomes across CAMERA and ADNI. **A.** Associations of brain-derived phosphorylated tau 231 (BDpTau231) with cognitive performance in CAMERA (left) and longitudinal changes in modified Preclinical Alzheimer’s Cognitive Composite (mPACC; middle) and Clinical Dementia Rating Sum of Boxes (CDR-SB; right) in ADNI. **B.** Associations of BDpTau231 with entorhinal cortical thickness in CAMERA (left) and AD-signature cortical thickness in ADNI (right). **C.** Associations of BDpTau231 with log-transformed white matter hyperintensity (WMH) volume in CAMERA (left) and ADNI (right). CAMERA models adjusted for age, sex and ethnicity, with additional adjustment for education and English as a first language for cognition and intracranial volume for WMH. ADNI longitudinal models adjusted for baseline age, sex, education and their interactions with time; MRI models adjusted for age, sex, imaging site and the interval between blood collection and MRI, with additional adjustment for intracranial volume for WMH.

**Extended Data Fig. 6.**
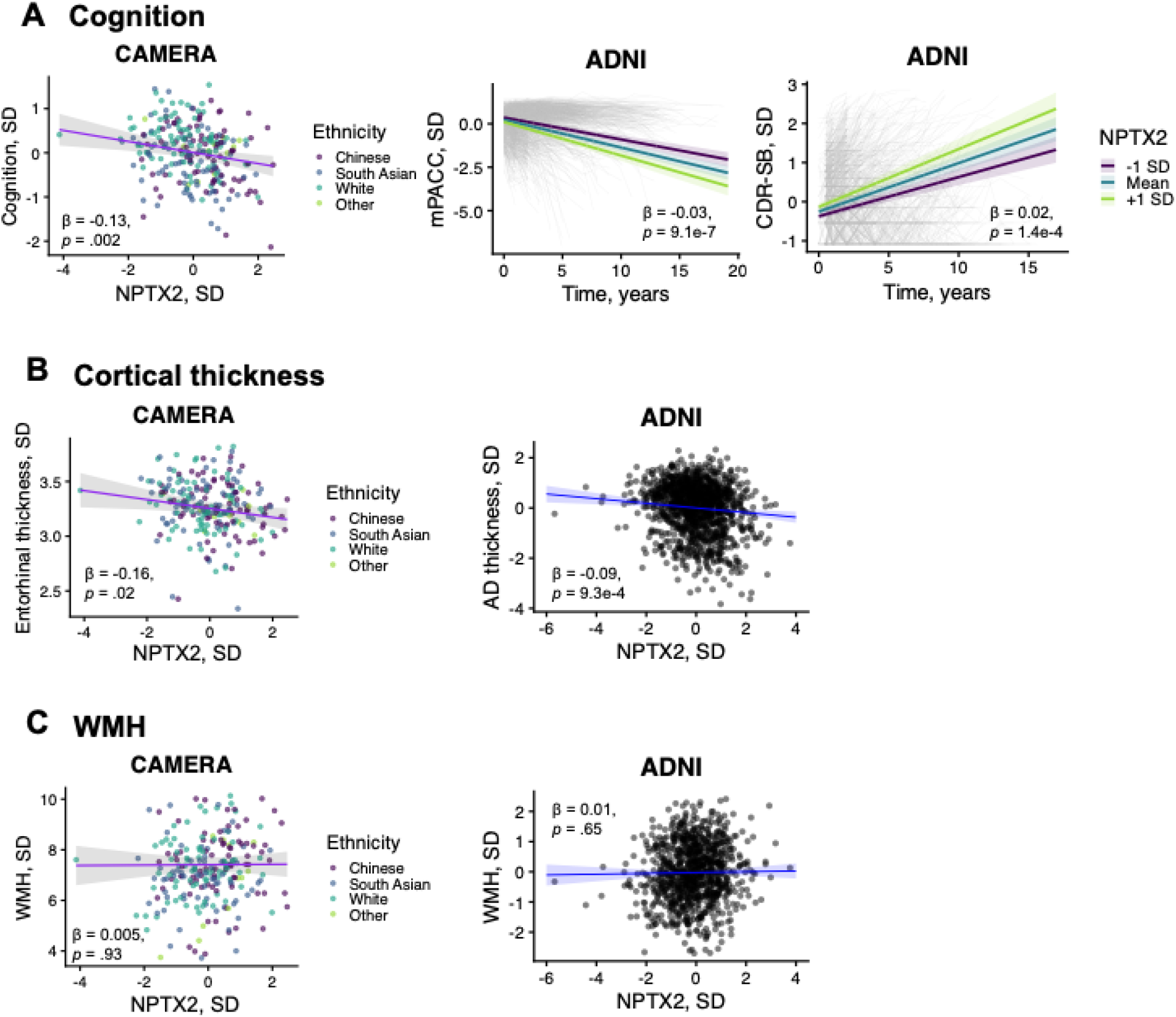
Associations of neuronal pentraxin 2 with cognitive and MRI outcomes across CAMERA and ADNI. **A.** Associations of neuronal pentraxin 2 (NPTX2) with cognitive performance in CAMERA (left) and longitudinal changes in modified Preclinical Alzheimer’s Cognitive Composite (mPACC; middle) and Clinical Dementia Rating Sum of Boxes (CDR-SB; right) in ADNI. **B.** Associations of NPTX2 with entorhinal cortical thickness in CAMERA (left) and AD-signature cortical thickness in ADNI (right). **C.** Associations of NPTX2 with log-transformed white matter hyperintensity (WMH) volume in CAMERA (left) and ADNI (right). CAMERA models adjusted for age, sex and ethnicity, with additional adjustment for education and English as a first language for cognition and intracranial volume for WMH. ADNI longitudinal models adjusted for baseline age, sex, education and their interactions with time; MRI models adjusted for age, sex, imaging site and the interval between blood collection and MRI, with additional adjustment for intracranial volume for WMH.

