## Supplemental figures for "Plasma proteomic markers of cognitive and brain aging in a multi-ethnic cohort"

**Figure S1. Associations of plasma NULISAseq proteins with precentral gyrus cortical thickness in CAMERA (as a control region), adjusted for age, sex, and ethnicity.**

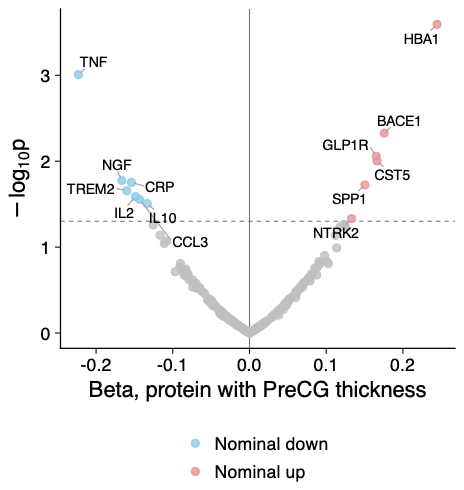

**Figure S2. Associations of plasma NULISAseq proteins with entorhinal cortical thickness in CAMERA (x-axis) and ADNI (y-axis), adjusted for age, sex, ethnicity (CAMERA only), as well as imaging site (ADNI only) and time between blood draw and MRI (ADNI only).**

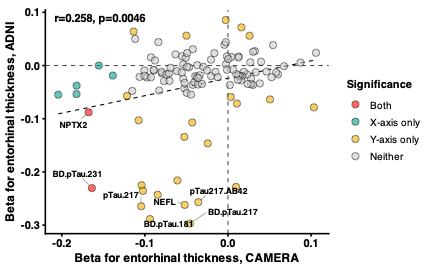

**Figure S3. Effect size comparison between main cognitive and brain proteomic models (x-axis) and models additionally adjusted for *APOE4* carriage (y-axis) in CAMERA.**

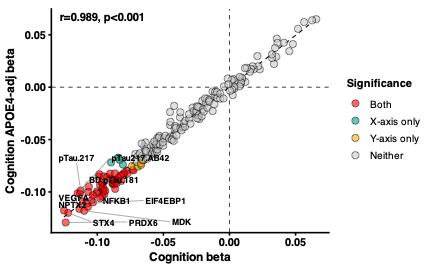

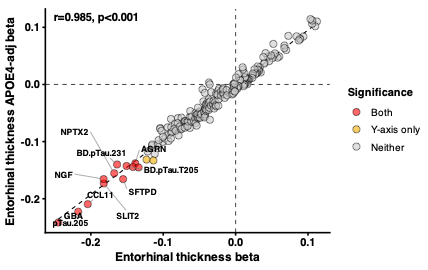

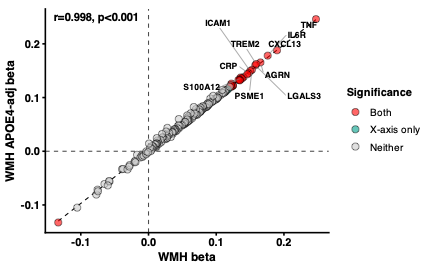

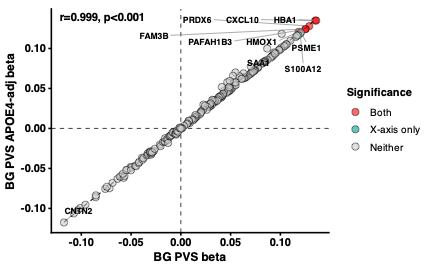

**Figure S4. Effect size comparison between main cardiometabolic proteomic models (x-axis) and models additionally adjusted for *APOE4* carriage (y-axis) in CAMERA.**

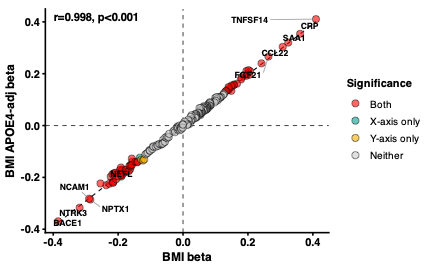

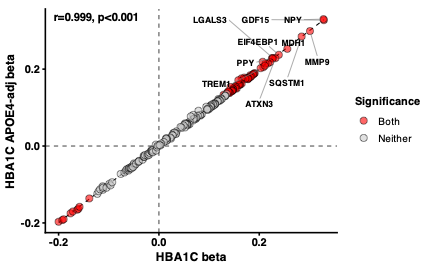

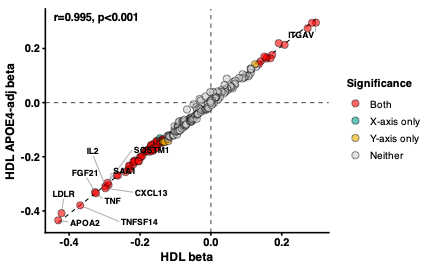

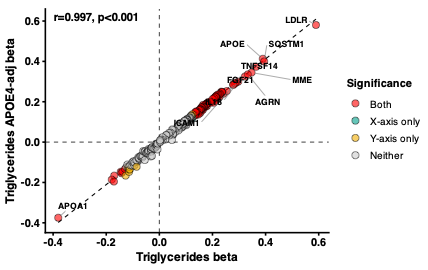

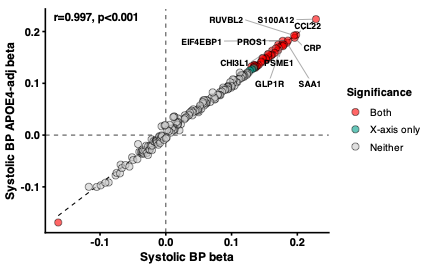

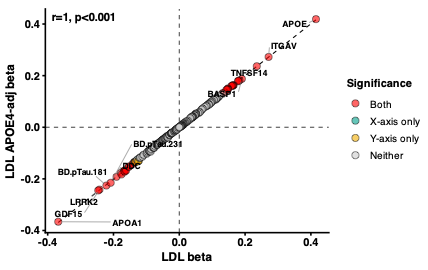

**Figure S5. Effect size comparison between main cardiometabolic proteomic models (x-axis) and models additionally adjusted for medication use (y-axis) in CAMERA.**

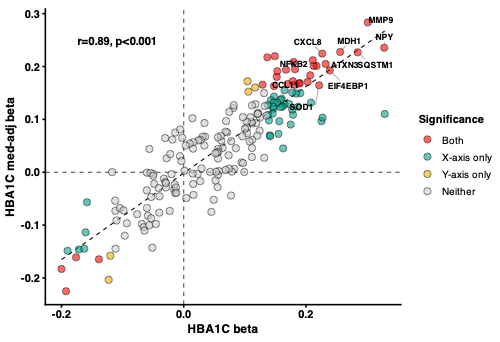

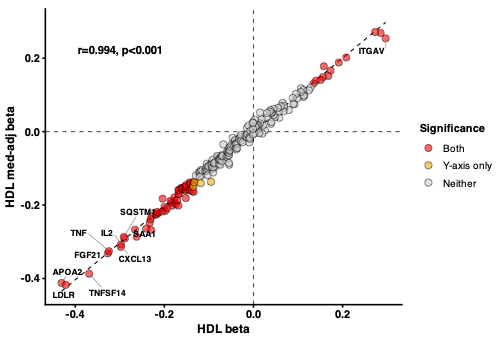

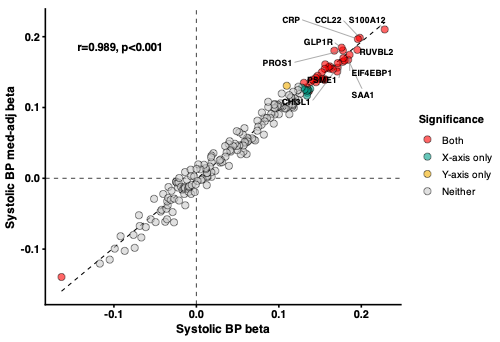

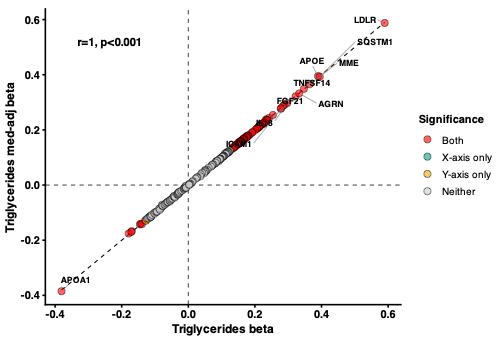

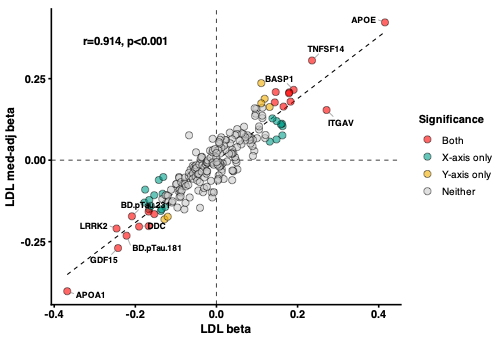

**Figure S6. Effect size comparison between main cardiometabolic proteomic models (x-axis) and models additionally adjusted for BMI (y-axis) in CAMERA.**

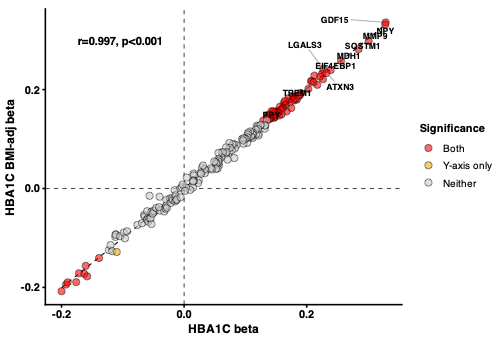

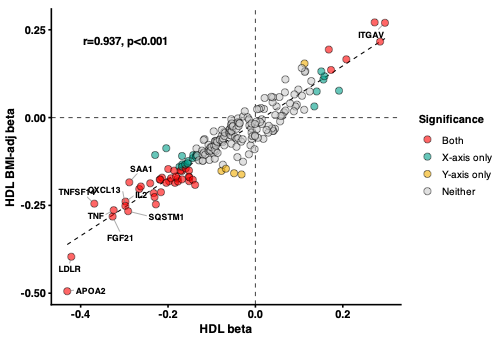

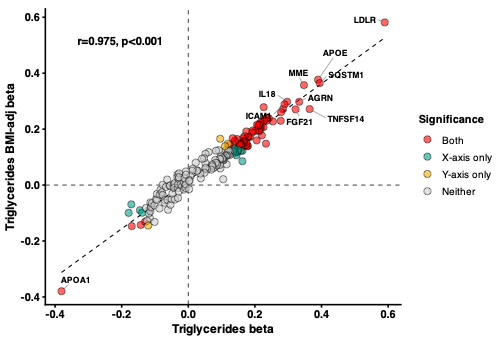

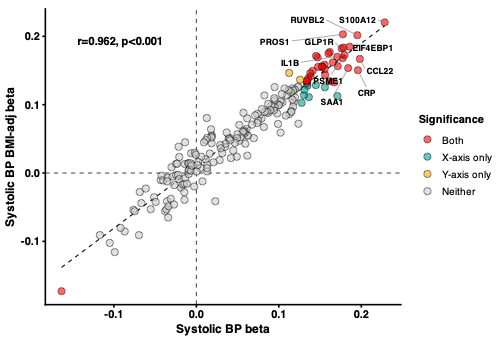

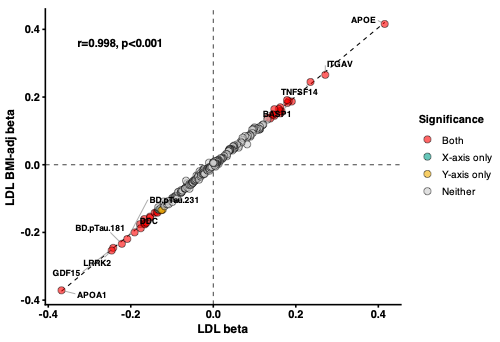

**Figure S7. Effect size comparison between main cognitive and brain proteomic models in ADNI in all participants (x-axis) and models excluding participants with dementia at baseline (y-axis).**

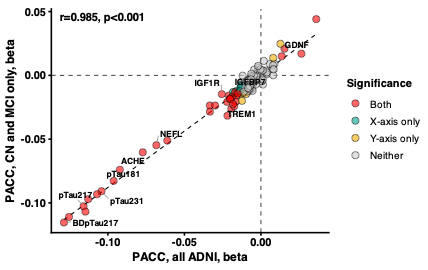

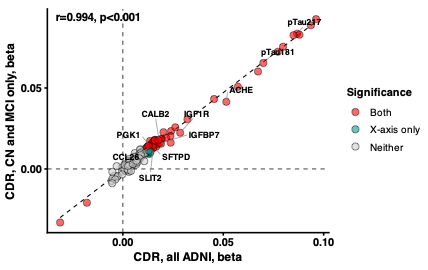

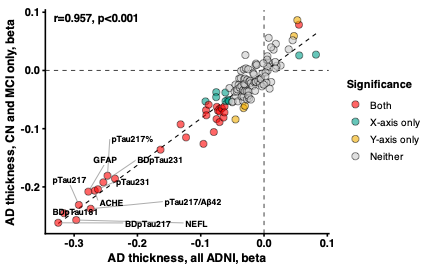

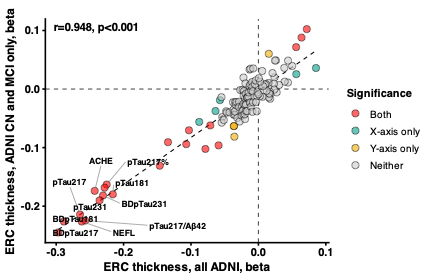

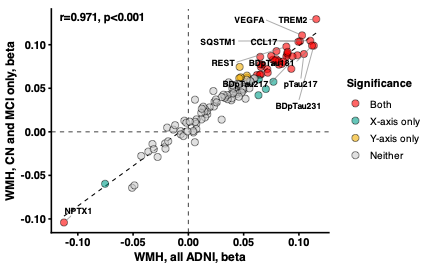

**Figure S8. Distribution of APOE4 protein in CAMERA.**

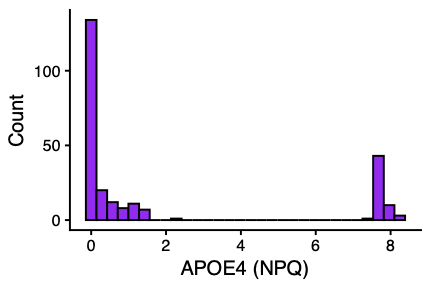
